# Integration of Deep Learning Annotations with Functional Genomics Improves Identification of Causal Alzheimer’s Disease Variants

**DOI:** 10.1101/2025.03.07.25323578

**Authors:** Chirag M Lakhani, Jui-Shan T. Lin, Anjali Das, Tatsuhiko Naito, Towfique Raj, David A. Knowles

## Abstract

Genetic variants associated with Alzheimer’s disease (AD) through genome-wide association studies (GWAS) are challenging to interpret because most lie in non-coding regions of the genome. Here, a method was developed that integrates deep learning variant effect prediction (DL-VEP) scores from Enformer, DeepSea, and ChromBPNet models with cell-type specific regulatory annotations to improve fine-mapping of causal AD variants. Using stratified linkage disequilibrium score regression, the largest proportion of heritability among brain cell types was found in microglia and the integration of Enformer microglia-based transcription factor DL-VEP scores and microglia ChromBPNet models further localizes the AD polygenic signal allowing for improved fine-mapping of causal AD variants. Functionally-informed fine-mapping using these annotations discovers 101 AD risk variants compared to 87 identified by statistical fine-mapping alone. Importantly, polygenic risk scores derived from these fine-mapped variants show improved cross-ancestry performance in both African American (AUC=0.532 vs. 0.521 for SUSIE) and Hispanic (AUC=0.552 vs. 0.535 for SUSIE) populations, while maintaining strong performance in European populations (AUC=0.620 vs. 0.615 for SUSIE). Through detailed analysis of the PICALM/EED locus, we demonstrate how our approach disentangles causal variants in regions of high linkage disequilibrium and predicts specific molecular mechanisms, such as disruption of PU.1 transcription factor binding, with the posterior inclusion probability of this variant increasing from 0.55 using SUSIE to 0.90 with our method. Our results provide a framework for leveraging deep learning annotations to improve identification of causal disease variants and enhance polygenic risk prediction across diverse populations.

## Introduction

Alzheimer’s disease (AD) is the most common form of dementia worldwide. In the US alone, it is believed that 7 million Americans are currently living with this disease^1^. Unfortunately, there are no known effective therapeutics for AD. One possible hope for an effective therapeutic would be to develop drug targets with strong genetic support. Recent analysis has shown that drug targets with human genetics support have double the chance of success^2^. Therefore, it is of utmost importance to better understand the genetic basis of AD. AD genetics is thought to exist on a spectrum^3^. On one end, studies of autosomally dominant AD implicate highly penetrant coding variants in APP, PSEN1, and PSEN2^4–6^. On the other end, GWAS of the more common late-onset sporadic AD^7–10^ has a more polygenic architecture, with many variants of smaller effect size throughout the genome.

Similar to other polygenic diseases, the majority of AD GWAS variants reside in non-coding regions of the genome. It is believed that most of these variants confer disease risk or protection through the modulation of gene expression or splicing. Using tools such as stratified linkage disequilibrium (LD) score regression (SLDSC)^11^, it has been shown that there is a strong enrichment of genomic signal in enhancer regions of microglia^12^. Further analysis^13–15^ has even mapped particular AD GWAS loci to specific enhancer regions in microglia, thereby implicating the causal cell type where these variants may act. However, understanding how variants in these loci contribute to AD requires identifying the causal variant and linking it to the causal gene. Various approaches such as Mendelian randomization^16^, transcriptome-wide association studies^17^, and colocalization methods^18^ are typically used to find the causal gene by looking for overlap in signal between a GWAS and eQTL study from the causal cell type. However, it is estimated that, on average, only 11% of trait heritability is mediated by cis-eQTLs^19^. In fact, there are distributional differences between variants implicated in GWAS compared to eQTL studies^20^. It has been observed that eQTL variants typically reside in more proximal promoter regions of the gene, whereas GWAS variants reside in more distal enhancer regions. Also, genes implicated in GWAS have stronger selective constraint compared to those implicated in eQTL studies. Recent approaches have shown that variants found to affect more proximal molecular events, such as chromatin accessibility, can explain a larger proportion of disease heritability^21,22^. Deep learning models that predict the output of functional genomics assays^23,24^ as a function of DNA sequence provide another approach for the prioritization of genetic variants that disrupt such proximal molecular events. Such models provide deep-learning based variant effect scores (DL-VEP), which predict the disruption of these events and are complimentary to collecting population level functional genomics data. In this analysis, these DL-VEP scores were utilized to better localize the AD GWAS polygenic signal and, in turn, use these DL-VEP annotations as priors^25^ for improved fine-mapping. We tested whether this approach can better uncover causal AD variants by testing whether polygenic risk scores (PRS) based on fine-mapped variants from a European-ancestry AD GWAS has better cross-ancestry portability. Finally, we show the utility of our integrative approach through an in-depth analysis of an AD GWAS locus in the PICALM/EED locus. This locus contains two variants in high LD, however, our method is able to prioritize one of the variants as being causal and also hypothesizes that this variant disrupts PU.1 transcription factor binding.

## Results

### Overview of Analysis

20,516 annotations were aggregated and generated from various data sources, including baseline annotations from Polyfun^25^, functional genomic profiles from Roadmap Epigenetics^26^ and brain cell types^12^, and deep learning models such as DeepSea^24^ and Enformer^23^ (**Methods and Supplementary Table 1**) to test for enrichment in AD GWAS^7–10^. For the functional genomics assays, we created annotations based on whether they fall into certain epigenetic regions of a cell type or whether they fall into a defined cis-regulatory region (CRE) based on a combination of epigenetic assays (**Methods**). For the deep learning models, we calculated DL-VEP scores (**Figure 1a**) using the functional genomics predictions given by DeepSea (2,002 assays) and Enformer (5,313 assays). A challenge with these deep learning models is that they were trained on publicly available datasets such as Roadmap Epigenetics^26^ and the ENCODE consortium^27^, which do not profile AD-relevant cell types such as microglia. To overcome this issue, we built upon an approach from Geijn et al.^28^ to generate annotations that combined cell-specific CRE annotations with cell-agnostic transcription factor (TF) binding DL-VEP scores (**Figure 1b**). Namely, we build cell-type specific annotations that only prioritize annotations within cell-type CRE regions with high TF DL-VEP scores. These annotations were meant to prioritize variants predicted to disrupt TF binding within the cell-type CRE region. Using all of the annotations, we implemented a two-stage process to find annotations that were relevant to AD genetics (**Figure 1c**). First, for each of the 20,329 non-baseline annotations, we ran SLDSC, including the baseline annotations as additional annotations. We found 381 annotations with nominally significant (p-value < 0.05) enrichment of heritability (e) and nominally significant *coefficient, representing an increase in per-SNP heritability conditioned on all other annotations^29^. In the second stage, in addition to the baseline annotations, we added two microglia annotations and then again selected annotations with nominally significant e and * with respect to the Bellenguez^8^ AD GWAS. These microglia annotations consisted of: 1) variants within microglia CRE regions (±500 bp flanking) and 2) variants within microglia CRE regions (±500 bp flanking) that also had a microglia TF DL-VEP z-score > 1. This resulted in 16 non-baseline annotations to be used for functionally-informed fine-mapping.

**Figure 1:**
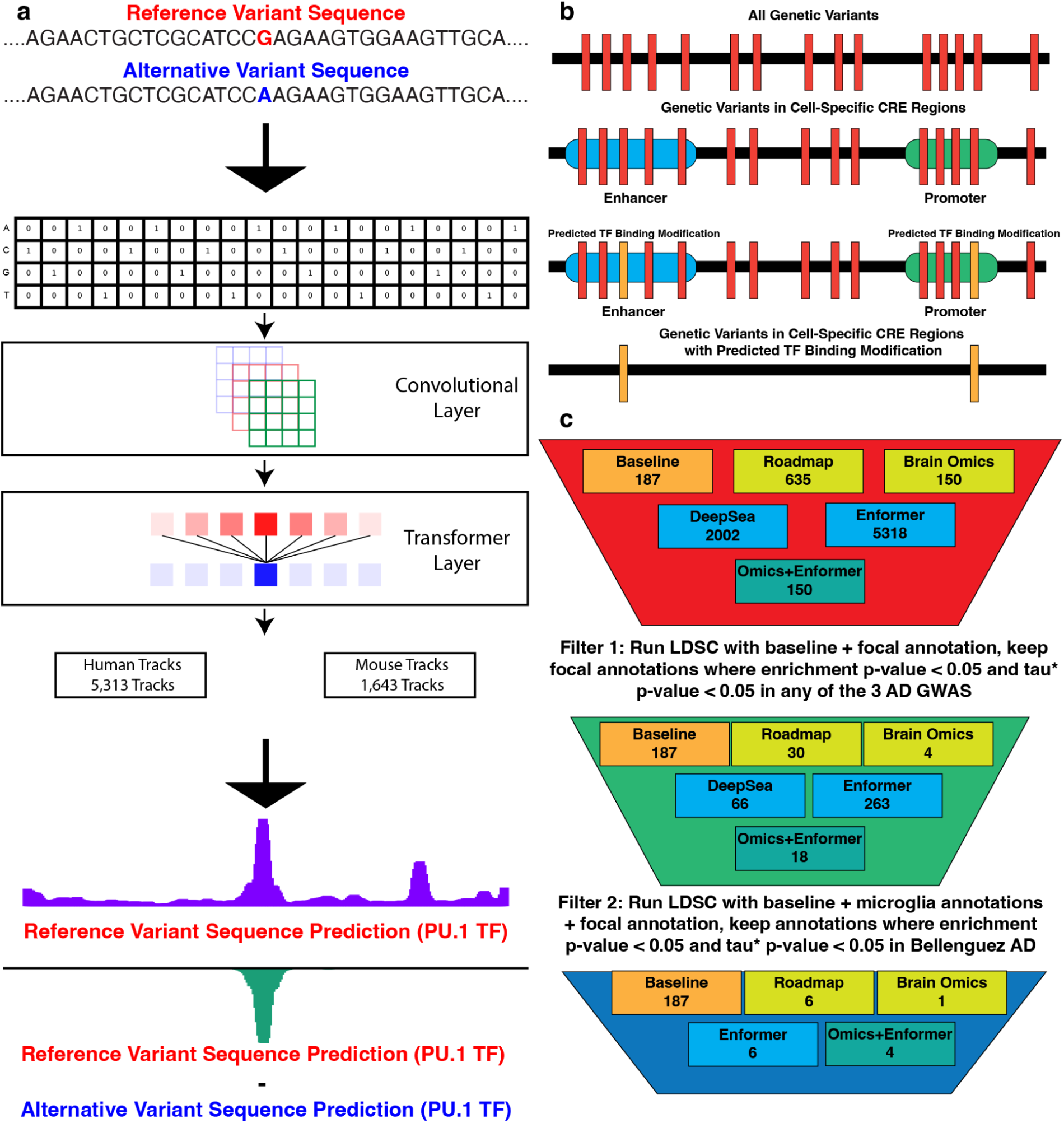
a) Deep learning variant effect prediction (DL-VEP) score calculation comparing reference and alternate allele sequences. b) Integration of cell-type specific cis-regulatory elements (CREs) with transcription factor disruption scores from deep learning models. c) Two-stage filtering process to identify functional annotations relevant to AD genetics.

We used the Bellenguez AD GWAS, consisting of 788,989 individuals of European ancestry, for our fine-mapping analysis and subsequent construction of AD PRS because of its large sample size and high genotyping density compared to older AD GWAS studies. We constructed a 20,848,201 variant LD reference panel from 17,192 individuals of European ancestry from the Alzheimer’s Disease Sequencing Project (ADSP)^30^ to mitigate LD reference panel mismatch bias known in PolyFun^25^ and to have maximal variant overlap with the Bellenguez GWAS (**Methods**). Once the final set of annotations were determined, we performed both statistical fine-mapping using SuSiE ^31^ and functionally-informed fine-mapping^25^ using three sets of functional annotations (Baseline, Baseline+Omics, and Baseline+Omics+DL) as priors to the SuSiE model. As an external validation of our fine-mapping methods, we constructed an AD PRS using the fine-mapped AD risk variants from the Bellenguez GWAS. We then scored individuals of African-American and Hispanic genetic ancestry from the ADSP data. It is well established that PRS derived from European-ancestry GWAS demonstrate diminished predictive accuracy when applied to non-European populations^32,33^, primarily due to three key factors: (1) differences in allele frequencies between populations affecting the statistical power to detect associations; (2) population-specific LD patterns causing European-identified tag SNPs to imperfectly capture true causal variants in non-European populations; and (3) potential differences in causal effect sizes across populations due to gene-environment interactions or epistasis that vary by ancestry. Recent analysis has shown that improved fine-mapping can aid in cross-ancestry portability of PRS by being less sensitive to varying LD^34^. Therefore, we constructed AD PRS based on fine-mapped SNPs, at different posterior inclusion probability (PIP) thresholds, found from statistical and functionally informed fine-mapping using all three sets of annotations. We applied these scores to the African-American and Hispanic populations from the ADSP cohort to determine the relative boost in AUC performance in those populations using the Baseline+Omics+DL annotations compared to other methods. Finally, we interrogated specific AD risk loci such as one in an intergenic region between the PICALM and EED genes. Of note, this locus contains two variants in very high LD making it difficult for standard statistical fine-mapping tools to determine the causal variant. However, when integrating the deep learning annotations, we have higher confidence in determining the causal variant. The deep learning models aid in determining the effects of this variant on transcription factor binding and prediction for the gene for which this variant is an eQTL. This demonstrates the ability of these deep learning tools to not only prioritize the causal variant but also aid in generating hypotheses about specific functional mechanisms for this variant.

### Microglia CRE regions contain the largest proportion of AD GWAS heritability

In AD genetics, it has been shown that enhancer/promoter regions of microglia have large enrichment of heritability^12,35^ demonstrating its importance as a cell type in the study of AD genetics. However, in order to further localize AD polygenic signal using deep learning models, it is important to quantify the total amount of AD heritability found in the CRE regions of each cell type. We calculated both the proportion of heritability (p) and e for three recent GWAS: Kunkle^7^ (KE), Wightman^9^ (WN) and Bellenguez^8^ (BZ). We found microglia to have both the highest p (p_KE_=0.604, SE=0.1880, p_WN_=0.261, SE=0.0464, p_BZ_=0.566, SE=0.1020) (**Figure 2b**) and highest e (e_KE_=12.30, SE=3.830, e_WN_=5.33, SE=0.949, e_BZ_=11.60, SE=2.080) (**Figure 2c**) among the 4 cell types (**Figure 2a**). Despite the high e, about 5% (**Figure 2a**) of genetic variants fall into microglia CRE regions, motivating the need for further localization of AD polygenic signal by prioritizing putatively causal variants in these loci with the potential to disrupt intermediate molecular phenotypes in these brain cell types.

**Figure 2:**
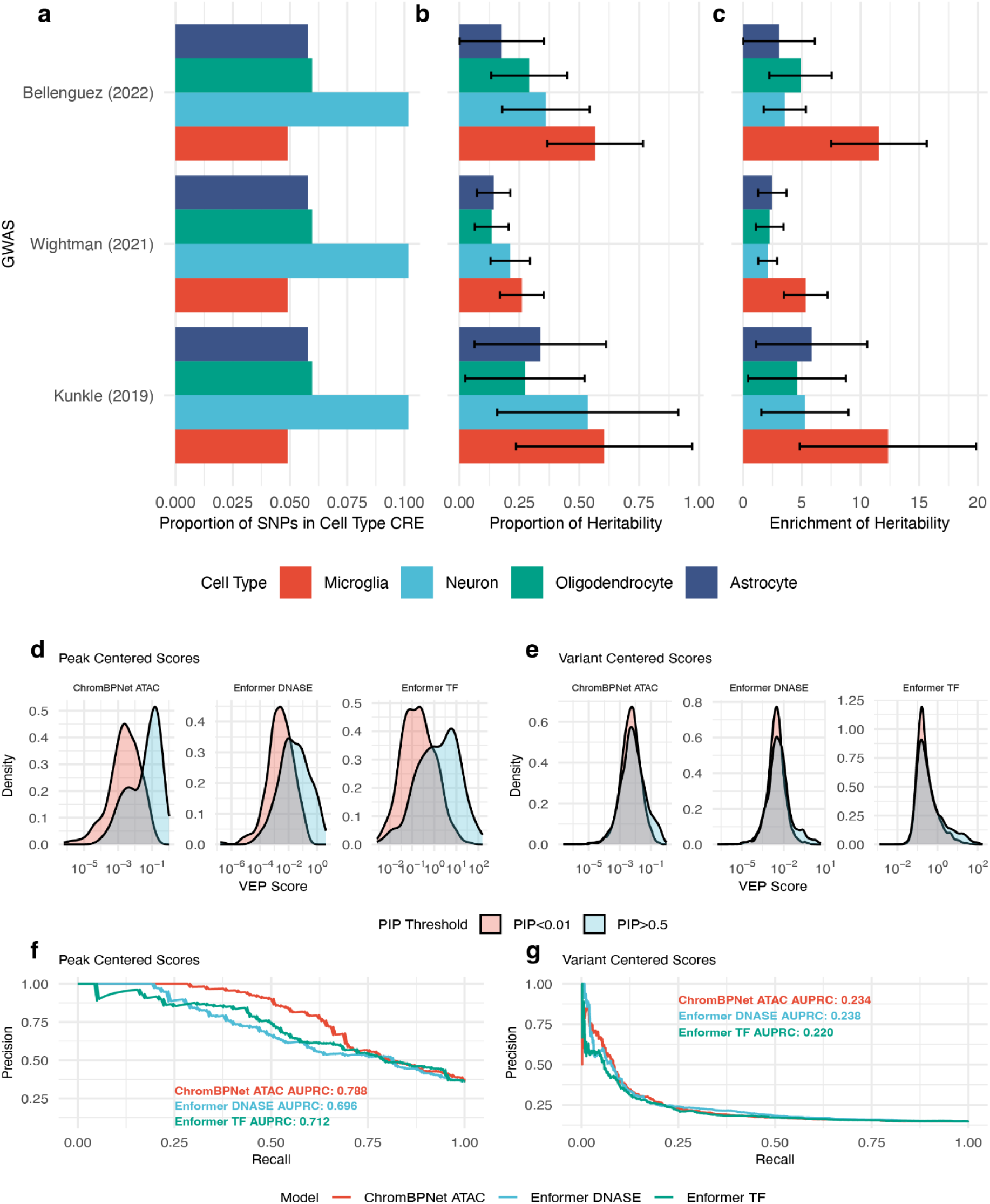
a) Distribution of genetic variants across CRE regions of four brain cell types. b) Proportion of heritability (p) captured by CRE regions across brain cell types for three AD GWAS studies. c) Enrichment of heritability (e) in CRE regions across brain cell types. d) Peak-centered analysis showing separation between causal and non-causal microglia caQTLs using DL-VEP scores. e) Variant-centered analysis demonstrating limited differentiation between causal and non-causal variants when not anchored to chromatin accessibility peaks.

We constructed DL-VEP scores from a ChromBPNet^36^ model trained on microglia ATAC-seq data, Enformer microglia TF DL-VEP scores (**Supplementary Table 2**) and Enformer DL-VEP score of DNASE from monoctyes (**Methods**). Using data from fine-mapped caQTLs from microglia^13^, we tested whether these scores could distinguish between causal and non-causal microglia caQTL within 1024 bp of a consensus chromatin accessibility peak (peak-centered) as well as whether it can distinguish between causal and non-causal caQTLs irregardless of the peak (variant-centered). We found separation in causal and non-causal caQTL VEP scores from the peak-centered analysis (**Figure 2d**) but little separation within the variant-centered analysis (**Figure 2e**). For the variant-centered approach, we found all three models had strong predictive performance, with the ChromBPNet DL-VEP scores having the highest (AUPRC=0.788), then Enformer microglia TF DL-VEP scores (AUPRC=0.712), and finally the Enformer DNASE DL-VEP score (AUPRC=0.696) (**Figure 2b**). However, for the variant-centered approach, the ChromBPNet (AUPRC=0.234), Enformer microglia TF (AUPRC=0.238), and Enformer DNASE (AUPRC=0.220) scores did not have strong predictive performance in separating causal from non-causal caQTLs in microglia. This provides evidence that an optimal strategy for utilizing deep learning VEP scores requires anchoring those predictions to regions with known CRE regions for the cell type of interest.

### Enrichment of heritability among functional annotations

In order to further localize the AD polygenic signal, we constructed binary cell-type specific annotations that prioritize genetic variants within cell-type CRE regions that have predicted TF disruption from the Enformer and ChromBPNet models (**Methods**). We z-score normalized the Enformer DL-VEP scores based on predictions from all ∼18 million variants within the UK Biobank reference panel. For each of the 4 brain cell types, we constructed three binary annotations (CRE+Enformer annotations) in which we selected variants within the cell type CRE region with predicted TF disruption of any TF (ATF) or cell-type specific TF (CTF) (Supplementary Table 2) at 3 different z-score thresholds (z-score > 1, 2, or 3). We found that enrichment of heritability using cell-type specific TFs had high enrichment of heritability while also maintaining close to the same proportion of heritability explained (**Supplementary Figure 1-4**) across all 3 AD GWAS. In particular, for the Bellenguez GWAS, the enrichment of heritability for annotations using DL-VEP scores in microglia is almost double that found for all variants in microglia CRE regions (e_BZ(CTF)_ = 22.2, SE = 4.54, e_BZ(ATF)_ = 13.8, SE = 2.68, e_BZ(CRE)_ = 11.6, SE = 2.08) while also maintaining close to the same proportion of heritability (p_BZ(CTF)_ = 0.508, SE = 0.104, p_BZ(ATF)_ = 0.543, SE = 0.105, p_BZ(CRE)_ = 0.566, SE = 0.102). For microglia, we found using a higher z-score threshold also increased e but proportionally dropped p (**Figure 3 and Supplementary Figure 5**), which motivated the use of the z-score = 1 threshold for downstream analysis. We also developed annotations where we added variants predicted to disrupt microglia chromatin accessibility based on ChromBPNet DL-VEP scores (**Methods**) to the microglia CRE+Enformer annotations (CRE+Enformer+ChromBPNet annotations). We found the microglia CRE+Enformer+ChromBPNet annotations did not meaningfully increase e or p when calculated using the UK Biobank reference panel (**Supplementary Figure 9**). However, when using the ADSP reference panel, p increased from p = 0.490 to p = 0.500 when using the ChromBPNet scores, resulting in p = 0.500 and e = 20.9 when using the microglia CRE+Enformer+ChromBPNet annotation compared to p = 0.599 and e = 12.6 with just the microglia CRE annotation (**Figure 3**). For subsequent analysis, we used the microglia CRE+Enformer+ChromBPNet annotation to maximize the amount of heritability captured by our deep learning annotation.

**Figure 3:**
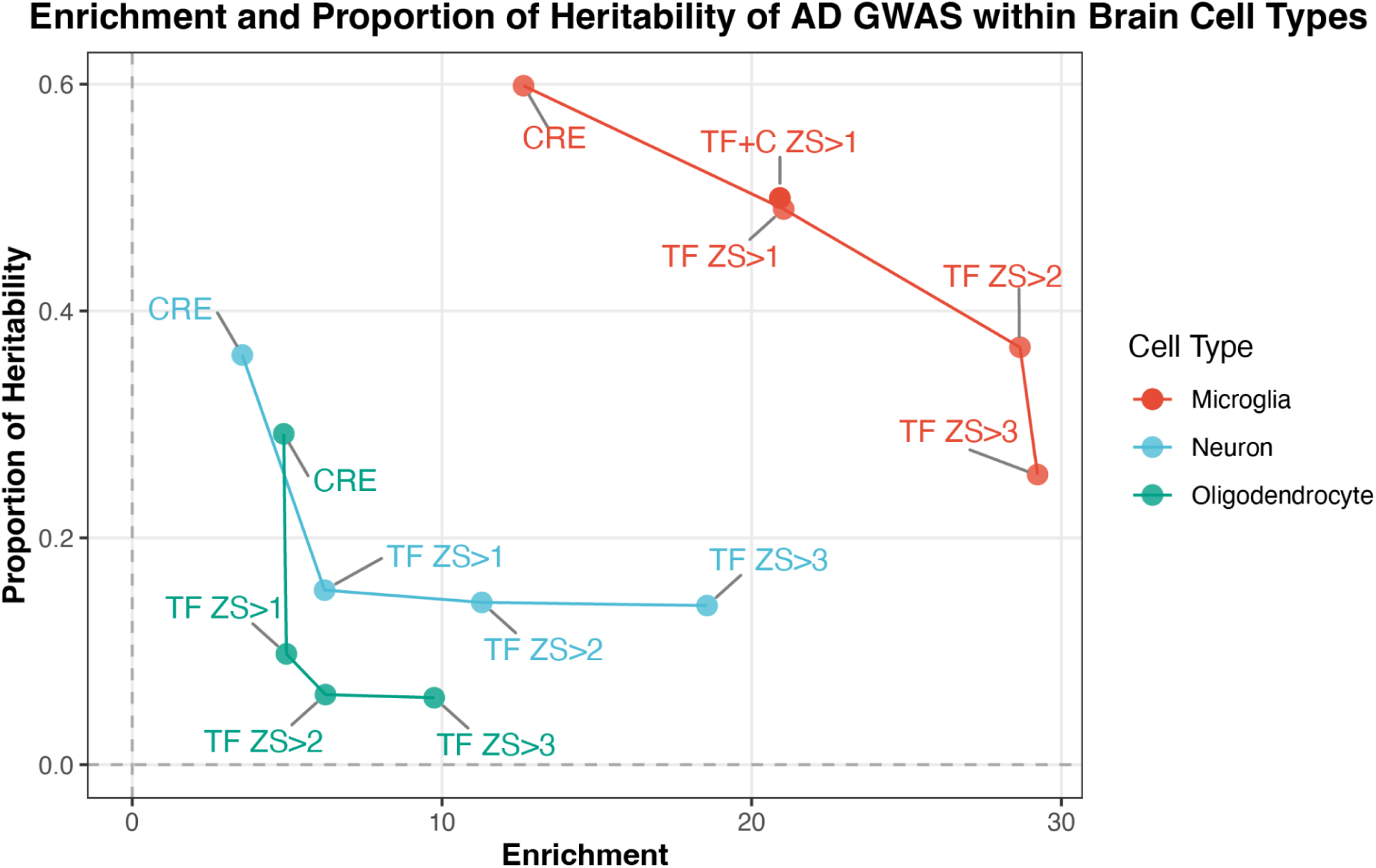
Comparison of heritability enrichment versus proportion of heritability for microglia, neuron, and oligodendrocytes. (astrocytes is not pictured due to the cell type having a negative proportion of heritability)

### Fine-Mapped Variants

As stated earlier, a large-scale search was conducted of functional annotations most relevant to AD GWAS through a two-step process resulting in a set of functional annotations with nominally significant * for the Bellenguez GWAS after including the baseline annotations and the two microglia annotations. In certain categories such as the Roadmap and Enformer annotations there were highly correlated cell type profiles, so for each assay the cell type with the highest p was selected and another cell type with the highest e value. The final list of annotations consisted of 6 Roadmap, 1 Brain Omics, 8 Enformer, and 2 Enformer + Omics annotations. We computed SLDSC using these annotations and found Enformer DL-VEP score for RNA Pol II disruption in spleen and microglia CRE+Enformer+ChromBPNet still had nominally significant * coefficients (**Figure 4a and Supplementary Table 3**). Using the SNPs from the Bellenguez GWAS, we performed statistical and functionally informed fine-mapping using three classes of annotations (Baseline, Baseline/Omics, and Baseline/Omics/DL) (**Supplementary Table 3**). For each category, we kept all nominally significant SNPs with PIP > 0.5. Functionally informed fine-mapping using the Baseline/Omics/DL annotations contained 77 credible sets and 24 single SNPs satisfying this criterion, whereas statistical fine-mapping resulted in 70 credible sets and 17 single SNPs (**Figure 4b**). Counterintuitively, the mean size of credible sets of variants fine-mapped using Baseline/Omics/DL was larger (n=1.77) compared to just SuSiE fine-mapping (n=1.51) (**Supplementary Figure 10**). Across all four fine-mapping approaches, we identified 138 nominally significant SNPs with PIP > 0.5, with 61 (44.2%) detected by all approaches, 21 (15.2%) unique to SuSiE, and 22 (15.9%) shared exclusively among the three functionally-informed methods (**Supplementary Figure 11**). As hypothesized, among the 3 non-SuSiE strategies, Baseline/Omics/DL annotations produced the largest number of newly fine-mapped SNPs (n=12, 9.02%) as well as the largest number of fine-mapped variants overall (n=101). Of these 101 variants, 37 were found in intronic regions, 34 in microglia CRE regions, 22 in neuron CRE regions, 19 still uncategorized, and 18 in coding regions (**Figure 4c**). Among variants found in a single cell type-specific CRE, microglia had the highest count (n=12), exceeding the number found exclusively in neurons, astrocytes, or oligodendrocytes.

**Figure 4:**
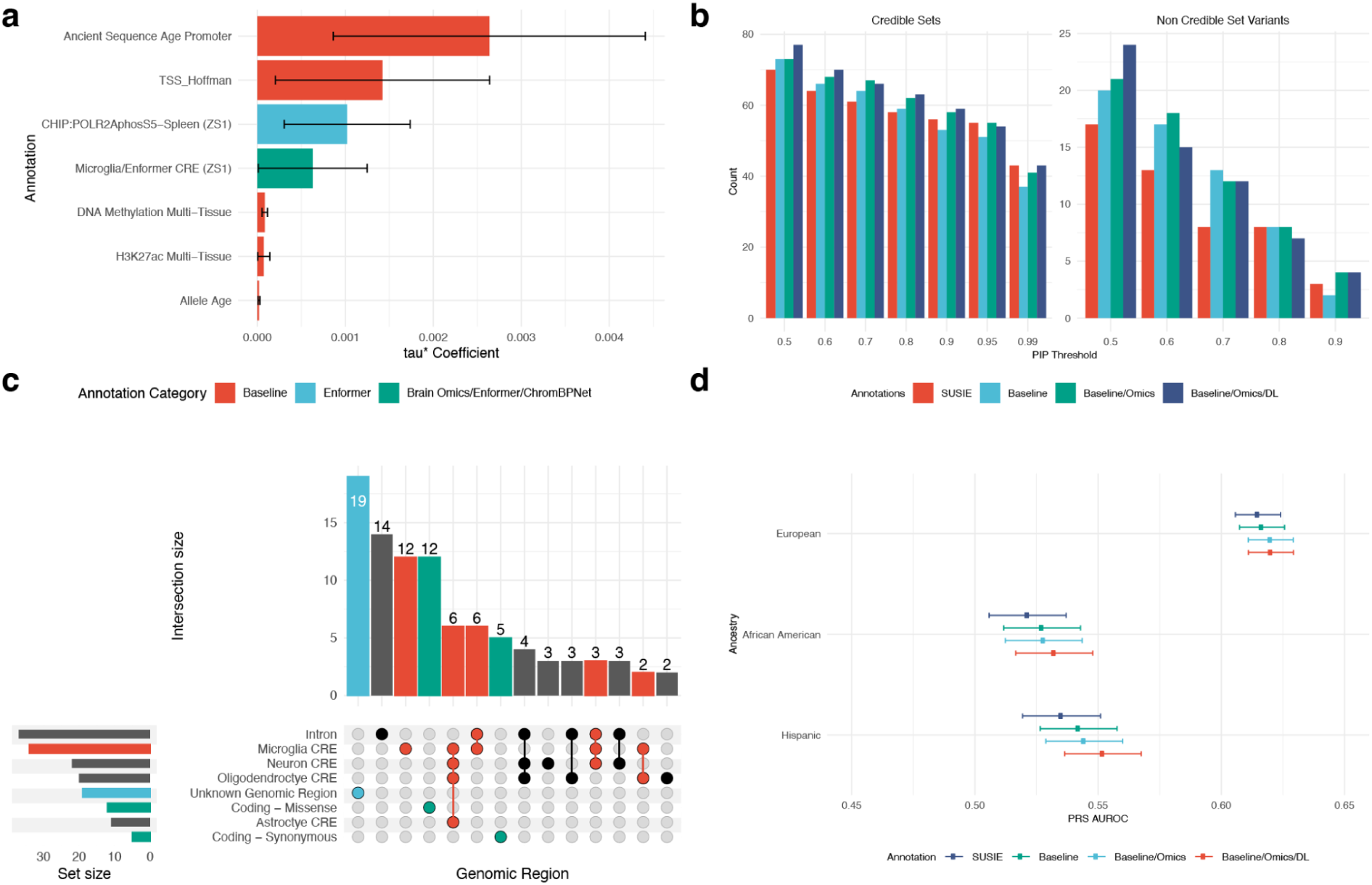
a) SLDSC analysis of final selected annotations with significant τ* coefficients. b) Counts of number of credible sets and single variants identified by statistical versus functionally-informed fine-mapping approaches. c) Genomic characterization of fine-mapped variants (PIP>0.5) using Baseline/Omics/DL annotations. d) PRS performance across European, African American, and Hispanic ancestries showing superior cross-ancestry performance with Baseline+Omics+DL annotations.

**Figure 5:**
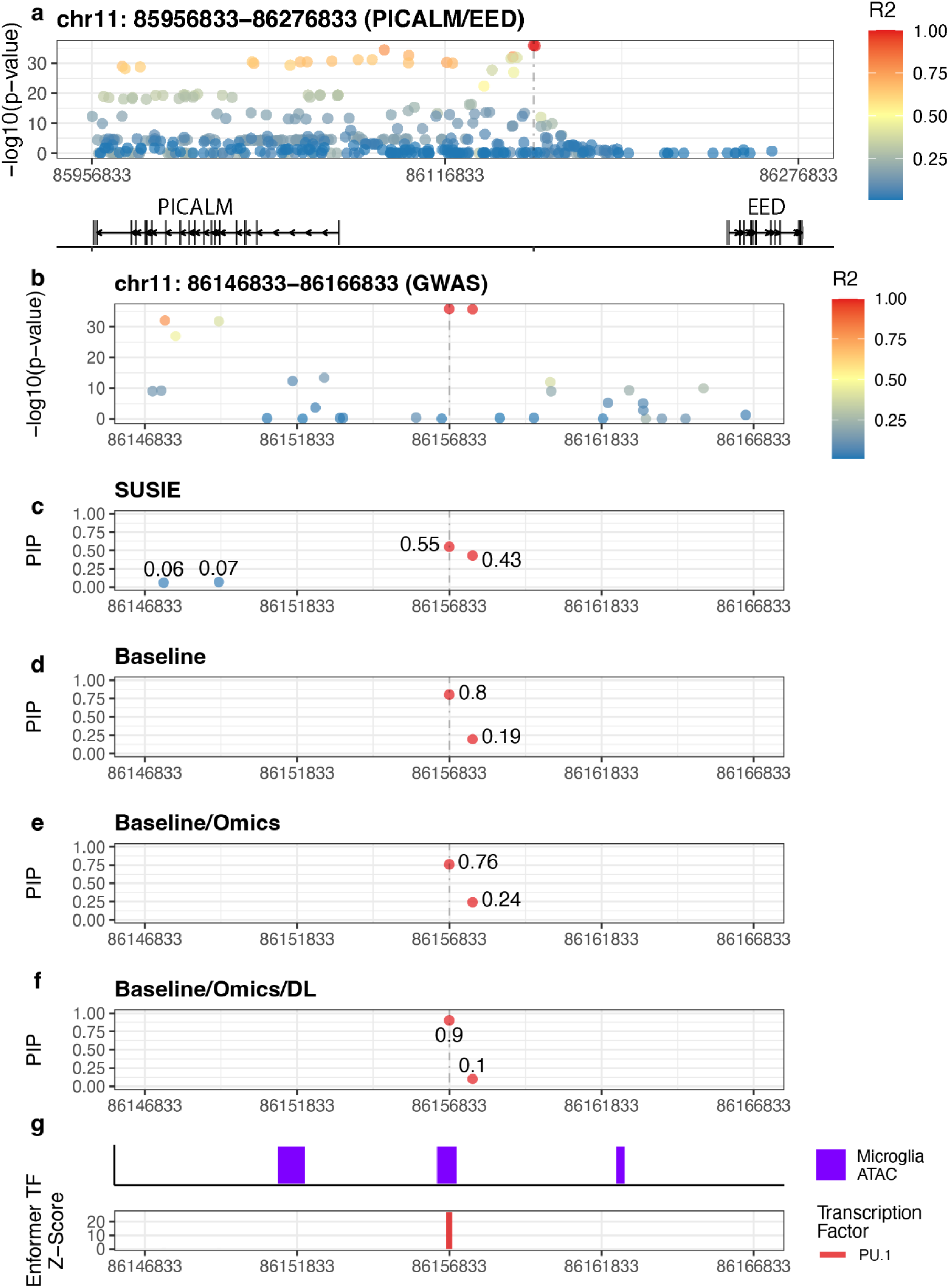
a) Regional association plot showing significant AD association signals. b) Zoomed-in GWAS association plot highlighting two highly correlated variants (r>0.90). c) Credible sets in locus using SuSie fine-mapping. d) Credible sets in locus using Baseline functionally-informed fine-mapping. e) Credible sets in locus using Baseline+Omics functionally-informed fine-mapping. f) Credible sets in locus using Baseline+Omics +DL functionally-informed fine-mapping. g) microglia ATAC-Seq peaks in locus and DL-VEP scores for PU.1 TF binding.

### Results of PRS in non European Ancestries

To evaluate whether deep learning annotations improve identification of causal AD variants, we tested if PRS constructed from fine-mapped SNPs show enhanced predictive performance in non-European populations. Using the latest ADSP data release, we analyzed 17,155 individuals of European ancestry, 5,947 of African-American ancestry, and 6,134 of Hispanic ancestry (**Table 1**). We constructed multiple PRS by selecting credible sets and individual SNPs above various PIP thresholds (0.50, 0.60, 0.70, 0.80, 0.90, 0.95, and 0.99) and scored each individual by multiplying the posterior effect size by the number of alternative alleles (**Methods**). For each population, we report the best-performing PRS score across all PIP thresholds. The Baseline+Omics+DL annotations yielded the highest AUC in both African-American and Hispanic ancestries (AFR: AUC=0.532, SE=0.00797, PIP=0.50; AMR: AUC=0.552, SE=0.00792, PIP=0.60; EUR: AUC=0.620, SE=0.00465, PIP=0.95), while SuSiE consistently produced the lowest performance (AFR: AUC=0.521, SE=0.00797, PIP=0.50; AMR: AUC=0.535, SE=0.00807, PIP=0.99; EUR: AUC=0.615, SE=0.00468, PIP=0.90) (**Figure 4d, Supplementary Table 3, Supplementary Figures 12-14**).

**Table 1:**
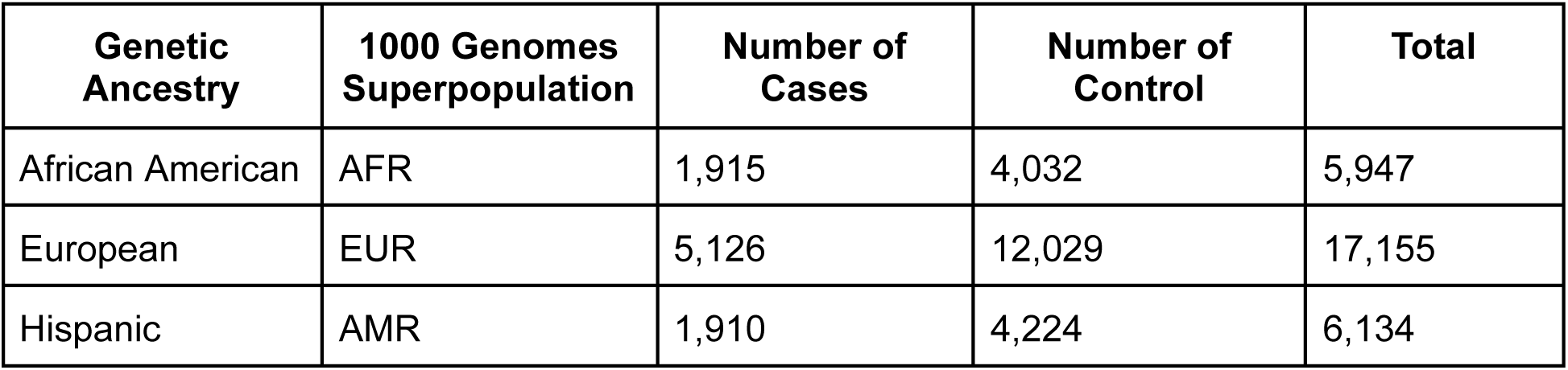
Demographic characteristics of the ADSP cohort by genetic ancestry, showing the number of cases and controls for each ancestry group.

| Genetic Ancestry | 1000 Genomes Superpopulation | Number of Cases | Number of Control | Total |
| --- | --- | --- | --- | --- |
| African American | AFR | 1,915 | 4,032 | 5,947 |
| European | EUR | 5,126 | 12,029 | 17,155 |
| Hispanic | AMR | 1,910 | 4,224 | 6,134 |

### Analysis of AD GWAS loci

Finally, we investigated individual AD GWAS loci to gain deeper insight into the fine-mapped variants and their predicted functional consequences. We identified 12 AD variants that localized exclusively to microglia CRE regions (**Supplementary Figures 15-38 and Supplementary Table 3**). Among these, we matched variants to genes using the Activity-by-Contact model^37^, where we found two variants matched to genes that differed from their nearest genes: rs2526377 matched to TSOAP1 rather than its nearest gene SCGB1C2, and rs4714447 matched to TREM1 rather than DUSP22. Nine of these 12 variants exhibited high microglia TF DL-VEP scores, with predicted binding disruption for transcription factors RUNX1, SMAD2, BACH1, PU.1, CREB1, IRF1, and SMAD5. Notably, CREB1 and IRF1 each showed predicted binding disruption at two distinct variant locations, suggesting these TFs may be particularly important in microglia-related AD pathogenesis. We also investigated a well known AD locus in the PICALM/EED locus (**Figure 4a**). It is of note that there are two variants in high LD (r > 0.90) (**Figure 4b**). SuSiE fine-mapping provided close to equal weight to both variants (PIP = 0.55 and PIP = 0.43) where as fine-mapping using Baseline+Omics+DL annotations assigns rs10792832 the highest weight (PIP = 0.90) (Figure 4f). Analysis of the microglia regulome^13^ has also shown that rs10792832 also falls into an open chromatin region of microglia (**Figure 4g**). Of note, this particular loci is also within a CRE region of oligodendrocytes, however gene-mapping via ABC scores shows this locus is linked to the CCDC81 in microglia but there is no match in oligodendrocytes. For this variant, we observed an exceptionally high microglia DL-VEP score for PU.1 (z-score = 26.8), strongly suggesting that this variant disrupts PU.1 transcription factor binding.

## Discussion

In this study, we show the utility of DL-VEP scores in uncovering causal AD GWAS variants. Following an approach from van de Geijn et al.^28^, we developed hybrid annotations which integrated cell-agnostic TF DL-VEP scores with AD-relevant functional genomics data which further localized the AD GWAS polygenic signal. We utilized these annotations, along with annotations using other DL-VEP scores, to develop priors which provided better fine-mapping of AD GWAS variants. We demonstrated that the fine-mapped variants found using this approach demonstrated better cross-ancestry portability than standard statistical fine-mapping as well as non deep learning based functional annotations. Finally, through a deep-dive into an AD locus in the PICALM/EED inter-genic region, we found our approach could not only identify the causal AD variant in a region of high LD but also hypothesize the variant disrupts PU.1 TF binding.

This approach was particularly successful in developing hybrid DL-VEP/functional genomic annotations for microglia but less successful in other cell types such as neurons or oligodendrocytes where some AD variants may act. This is the case because our annotations look for genome-wide enrichment is strongest in microglia. It is also possible that the de novo motif analysis performed by Nott et al.^12^ is insufficient in identifying all of the important cell-type specific TFs. In future analysis, we plan to develop annotations which are both cell type and pathway specific. This approach has the advantage of better localizing the AD GWAS signal to not only the relevant cell type but also the cell process that may be disregulated in that cell type. For example, it is believed there is enrichment of AD GWAS signal for variants which affect genes involved in microglial efferocytosis^38^ but there are also AD genes involved in cholesterol metabolism, amyloid plaque, and neurofibriarlary tangle formation^3^. By restricting to annotations by both cell type and pathway we can uncover cell-type specific processes implicated in AD. It is understood that cell-type specific TF DNA binding is influenced by many other factors^39^ beyond just the DNA motif. One approach would be to use neural network interpretation methods trained on cell-type specific ATAC-Seq^36^ data to learn to identify TFs with more complicated binding syntax. Rather than use large models trained on thousands of assays, another approach would be to use TF DL-VEP scores from models specifically designed for TF assays such as BPNet^40^; these models explicitly incorporate CHIP control data to minimize enzyme bias. In future analysis, we will analyze each of these strategies in order to find the optimal approach for ascertaining cell-type specific TF DL-VEP scores for analysis of AD GWAS. We also note that our analysis was restricted to SNPs, primarily due to the lack of robust methods for constructing DL-VEP scores for indels. A future goal would be to also construct DL-VEP for indels in order to incorporate those variants into our analysis.

In our PRS analysis, while our deep learning informed approach performed best, there was still a drop-off in predictive accuracy when tested in the European population compared to both the African-American and Hispanic population. Some of this can be attributed to the choice of using the Bellenguez GWAS^8^ for fine-mapping. We chose this GWAS primarily because it is the most recent AD GWAS and it also has the highest genotyping density. However, the estimated SNP-heritability based on SLDC is only h^2^=0.0596 (SE=0.0067) compared to the older Kunkle GWAS, which has h2=0.1286 (SE=0.0406). This is explained by the fact that newer AD GWAS include AD proxy cases, which seem to dilute genetic heritability^41^. We anticipate utilizing our approach on WGS data from the ADSP cohort, which will allow for better fine-mapping of AD genetic variants due to having both high genotype density and stricter inclusion criteria for cases and controls. We also anticipate applying our approach to the multiple ancestries in the ADSP data, which will give us a robust set of annotations for functionally-informed fine-mapping. We would like to emphasize that our AD PRS are not meant to be clinically useful but only as an internal validation for whether our methods are better at capturing causal variants. We anticipate that our methodology could eventually lead to robust clinically-relevant AD PRS in the future. In this work, we provided a framework for the integration DL-VEP and functional genomics annotations for better fine-mapping of AD variants. Through better fine-mapping, we will be able to both determine causal variants driving AD GWAS risk and potentially better aggregate these findings to have a deeper understanding of the biological processes driving AD risk. Recent analysis has also shown that such integrative approaches can also aid with finding genes whose rare non-coding variant burden is associated with AD^42^. We believe such integrative approaches will aid in our understanding of the genetic architecture of AD and hopefully more successful therapeutics in the future.

## Methods

### Genetic Data

We utilized summary statistics from four recent Alzheimer’s disease GWAS, namely, Kunkle (2019)^7^, Jansen (2019)^10^, Wightman (2021)^9^, and Bellenguez (2022)^8^, along with the UK Biobank reference panel for the stratified LD-score regression analysis. For the functionally-informed fine-mapping analysis, we constructed an LD reference panel using 17,192 European samples and 20,848,201 variants (MAF > 0.005) from the Alzheimer’s Disease Sequencing Project (ADSP)^30^ whole genome sequencing dataset. We calculated LD scores for all annotations using this LD reference panel and subsequently used this reference panel for functionally informed fine-mapping with the Bellenguez (2022) GWAS.

We initially processed the data from the Alzheimer’s Disease Sequencing Project (ADSP)^30^ and performed the following quality control steps:

1. Removed SNPs with more than 10% missingness across all individuals.
2. Excluded individuals with greater than 10% missingness across variants.
3. Filtered out individuals with identity-by-descent (IBD) kingship relationships.
4. Excluded individuals lacking an Alzheimer’s disease diagnosis and age information.
5. Removed cases and controls under 65 years of age.

After the quality control steps, for our PRS analysis, we extracted individuals of African American, European, and Hispanic ancestry from the ADSP dataset. We utilized somalier^43^ to predict whether an individual belonged to one of three 1000 Genomes superpopulations. Individuals predicted to be in one of the 3 super populations (**Table 1)** were kept for further analysis. We remove all individuals except those controls under the age of 65 at the time of their last follow-up visit.

### Brain cell type CRE definition

We utilized the three epigenetic assays (ATAC-Seq, H3K27ac histone, and H3K4me3 histone) from Nott et al.^12^ and defined a brain cell type CRE to be the union of all ATAC-Seq, H3K27ac, and H3K4me3 peaks plus an extra 500 bp flanking sequence of these peaks.

### Deep Learning Variant Prediction Scores (DL-VEP)

For the calculation of deep learning variant effect prediction (DL-VEP) scores, we employed model-specific approaches to quantify the impact of genetic variants on predicted functional genomic outcomes. For Enformer predictions, we computed the difference between reference and alternate allele sequences across the middle 32 bins, corresponding to a 4,096 bp region centered on each variant, for all non-CAGE-Seq assays. With DeepSea, DL-VEP scores represent the difference in predicted binding probabilities between reference and alternate sequences. For ChromBPNet, which offers base-resolution predictions of chromatin accessibility, we calculated the difference between reference and alternate allele sequences across the middle 1,000 bp region. Through empirical testing against known chromatin accessibility QTLs (caQTLs), we determined that a ChromBPNet difference score threshold of 0.15 effectively separated caQTLs from non-caQTLs based on data from Kosoy et al.^13^ Based on this finding, we binarized ChromBPNet DL-VEP scores, assigning a value of 1 to variants with scores above 0.15, and 0 otherwise. To enable cross-assay comparisons and establish meaningful thresholds for DeepSea and Enformer, we z-score normalized their DL-VEP values using the distribution of approximately 18 million variants from the UK Biobank reference panel. This normalization procedure allowed us to identify variants with unusually large predicted effects relative to the genome-wide background distribution.

### Peak-centered versus Variant-centered Analysis of microglia caQTL

Two complementary approaches were employed to evaluate the performance of DL-VEP scores in distinguishing causal from non-causal microglia caQTLs. In the peak-centered approach, caQTLs and matched control variants were restricted to those located within 1024 bp of a consensus chromatin accessibility peak^13^. DL-VEP scores were calculated as the difference between predictions for reference and alternate allele sequences across the middle 1000 bp region centered on the peak. This approach was designed to test discrimination power in regions with established regulatory potential. In contrast, the variant-centered approach included all caQTLs regardless of their proximity to consensus peaks, with DL-VEP scores calculated for the middle 1000 bp region centered directly on each variant. This comparison allowed for assessment of whether the predictive capacity of deep learning models is dependent on the anchoring of variants to known regulatory regions. The superior performance observed in the peak-centered analysis (AUPRC=0.788 for ChromBPNet) compared to the variant-centered approach (AUPRC=0.234) underscores the importance of integrating variant effect predictions with cell-type specific regulatory annotations.

### Annotations

We compiled 20,516 variant annotations from the baseline-LF model^29,44,45^ and various functional genomics^12,26^ and deep learning^23,24,46^ data sources to determine the annotations most relevant to Alzheimer’s disease. For the functional genomics annotations, we ascertained bed files for DNase, H3K27ac, H3K7me3, H3K4me1, H3K4me3 assays for 127 cell types from the Roadmap^26^ consortium. For each assay-cell type pair, we generated a binary annotation based on whether a variant falls in or out of the assay peak region. We also ascertained binary annotations for 4 brain cell types (astroctyes, microglia, neuron, and oligodendrocytes) from Nott et al.^12^ based on whether a variant falls into a cell-type specific predicted cis-regulatory region (CRE) for each of these 4 cell types (**Supplementary Methods**). For the deep learning annotations, we calculated deep learning variant effect prediction (DL-VEP) scores for functional genomic assays from DeepSea^24,46^ (2002 assays), Enformer^23^ (4506 assays) deep learning model, and a ChromBPNet model (**Supplementary Methods**). To compare VEP scores across assays, we z-score normalized the VEP score for each assay using the variants in the UK Biobank reference panel provided by PolyFun^25^. The UK Biobank variant scaling was also applied to deep learning annotations created using the ADSP reference panel. For each VEP score, we generated 3 binary annotations based on whether the variant has a VEP z-score value greater than 1,2, or 3. From stratified LD Score Regression (SLDSC), for each annotation, we computed three quantities of interest:

1. Proportion of heritability (p) - SNP-heritability captured by variants in an annotation divided by the total SNP-heritability for the trait
2. Enrichment of heritability (e) - Proportion of heritability captured by the annotation (p) divided by the percentage of variants found in the annotation
3. Tau* coefficient ( * - proportionate change in per-SNP heritability per 1 standard deviation change in annotation value. This measure captures the effect unique to this annotation after conditioning on all other annotations.

We used a two-step process to determine the most relevant non-baseline annotations for AD GWAS.

1. For each non-baseline annotation (Supplementary Table 1), we ran stratified LD score regression. We included all baseline annotations plus the annotation of interest (focal annotation) across three AD GWAS^7–9^. We retained any focal annotation with a nominally significant (p-value < 0.05) e and nominally significant * among the three AD GWAS.
2. Next, in addition to the baseline annotations, we added two additional microglia-specific annotations in order to remove annotations that are capturing microglia signal:

1. Variants falling into a microglia CRE region (enhancer + promoter + 500 bp flanking)
2. Variants falling into a microglia CRE region (enhancer + promoter + 500 bp flanking) that also have an Enformer TF VEP z-score > 1 (among any microglia TFs).

We ran SLDSC and then identified focal annotations with nominally significant e and * (**Figure 1b**) for the Bellenguez GWAS.^8^ For the Roadmap and Enformer annotations there were highly correlated assays (e.g. similar cell types) so we added an additional filtering step. For the Roadmap and Enformer annotations, for each assay, highest proportion of heritability and another with highest enrichment of heritability. For the Enformer + Brain Omics, all three of the microglia annotations (one for each z-score threshold) passed our filtering criterion. We kept the microglia annotation with z-score threshold > 1 in order to capture the largest proportion of heritability.

### Functional fine-mapping

We took the 17,192 EUR samples with 7 million SNPs to build our LD reference panel. We used 3 MB LD blocks with a 1 MB overlap, then aggregated the result using the most centric LD block, as described in the PolyFun paper. Subsequently, we conducted functional fine-mapping with 3 different sets of annotations which are baseline, baseline+omics, baseline+omics+dl, and compared these results with SuSiE (fine-mapping without annotation).

### PIP value thresholding

After completing the fine-mapping step, we obtained the posterior inclusion probability (PIP), effect size (beta), and credible sets. Each credible set includes an index that indicates the order of the credible sets found within a specific window, ranging from 0 to the maximum number of credible sets pre-set during fine-mapping (which we set to 10). An index of 0 indicates that those SNPs were not included in any credible set for that block and are therefore treated as noise in our analysis. We remove all such SNPs and apply thresholds on PIP values for the remaining SNPs during PRS calculations to ensure that at least one SNP in the credible set exceeds the specified threshold. The number of SNPs passing each threshold for different sets of annotations can be found in supplement figure.

### Calculation of Polygenic Risk Score

We used PLINK 1.9 to calculate individual PRS, by multiplying the posterior effect size from fine-mapping by the number of alternate alleles. To evaluate PRS performance, we fit a logistic regression model where we predict AD case/control status as a function of individual PRS score. We assessed model performance by calculating the area under the receiver operating characteristic curve (auROC) and R^2^. We also calculated the standard error of the auROC metric using the DeLong test^47^.

## Supplementary Information

Supplementary document - Supplementary Figures 1-38; Supplementary Tables 1,2,5 Supplementary table 3 - final functional annotation list used for fine-mapping Supplementary table 4 - fine-mapped AD GWAS variant list

## Declarations

### Ethics Approval and Consent

Not Applicable

## Data Availability

This paper uses the Alzheimer's Disease Sequence Project Release 5 Whole Genome Sequencing data and Alzheimer's Disease phenotype data. Summary statistics for Alzheimer's disease GWAS are publicly available through the GWAS catalog. Ascension ID: GCST007511, GCST90044699, GCST90027158.

https://www.ebi.ac.uk/gwas/studies/GCST007511

https://www.ebi.ac.uk/gwas/studies/GCST90044699

https://www.ebi.ac.uk/gwas/studies/GCST90027158

## Acknowledgements

Research reported in this paper was supported by the Alzheimer’s Disease Sequencing Project of the National Institutes of Health under award number U01 AG068880-02. The content is solely the responsibility of the authors and does not necessarily represent the official views of the National Institutes of Health.

This work was also supported in part through the computational and data resources and staff expertise provided by Scientific Computing and Data at the Icahn School of Medicine at Mount Sinai and supported by the Clinical and Translational Science Awards (CTSA) grant UL1TR004419 from the National Center for Advancing Translational Sciences. Research reported in this publication was also supported by the Office of Research Infrastructure of the National Institutes of Health under award number S10OD026880 and S10OD030463. The content is solely the responsibility of the authors and does not necessarily represent the official views of the National Institutes of Health.

## Code Availability

Code and details for running analysis will be available on GitHub: https://github.com/daklab/AD_DLVEP upon journal publication

