## Supplementary Information for "Integration of Deep Learning Annotations with Functional Genomics Improves Identification of Causal Alzheimer’s Disease Variants"

#### Supplementary Figures

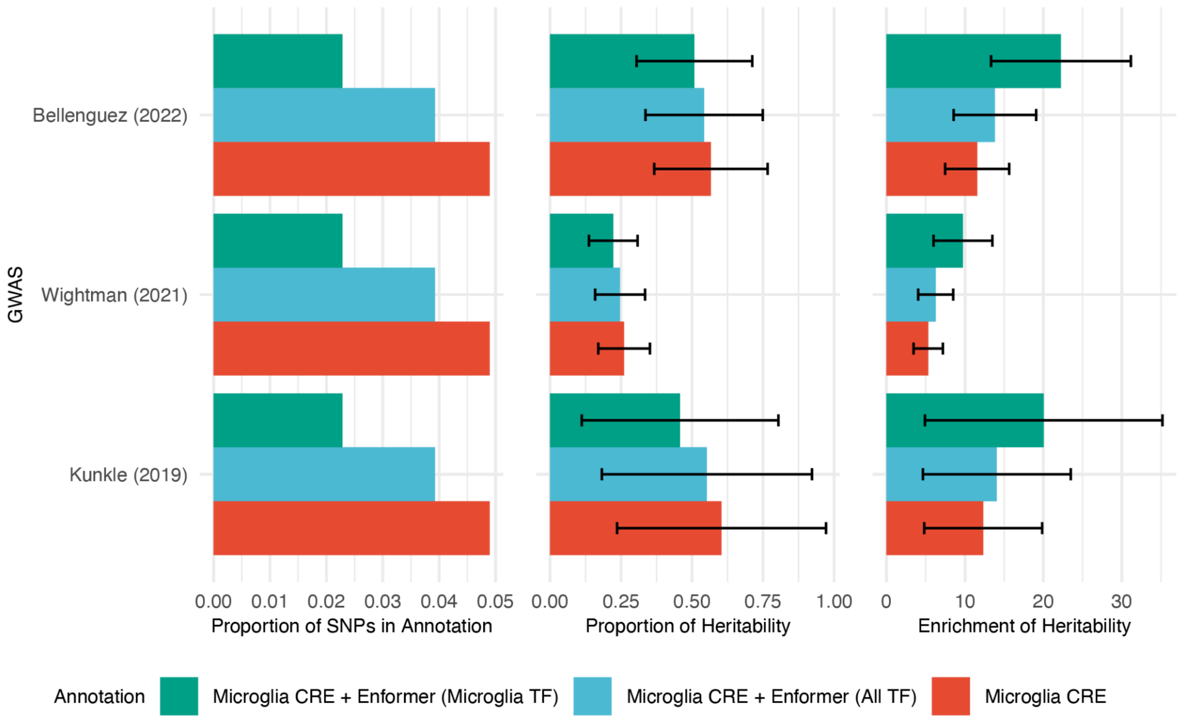


Supplementary Figure 1: Comparison of enrichment and proportion of heritability for cell-type specific TF (CTF) versus all TF (ATF) annotations at different z-score thresholds for microglia for AD GWAS.

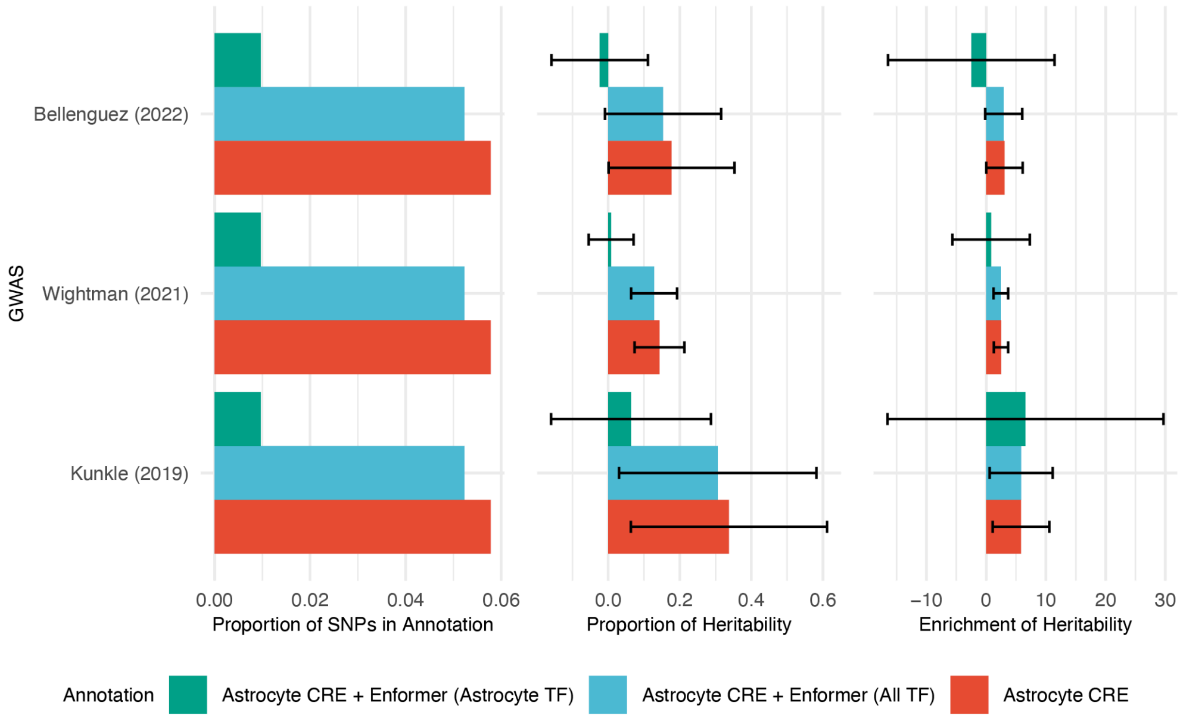


Supplementary Figure 2: Comparison of enrichment and proportion of heritability for cell-type specific TF (CTF) versus all TF (ATF) annotations at different z-score thresholds for astrocytes for AD GWAS.

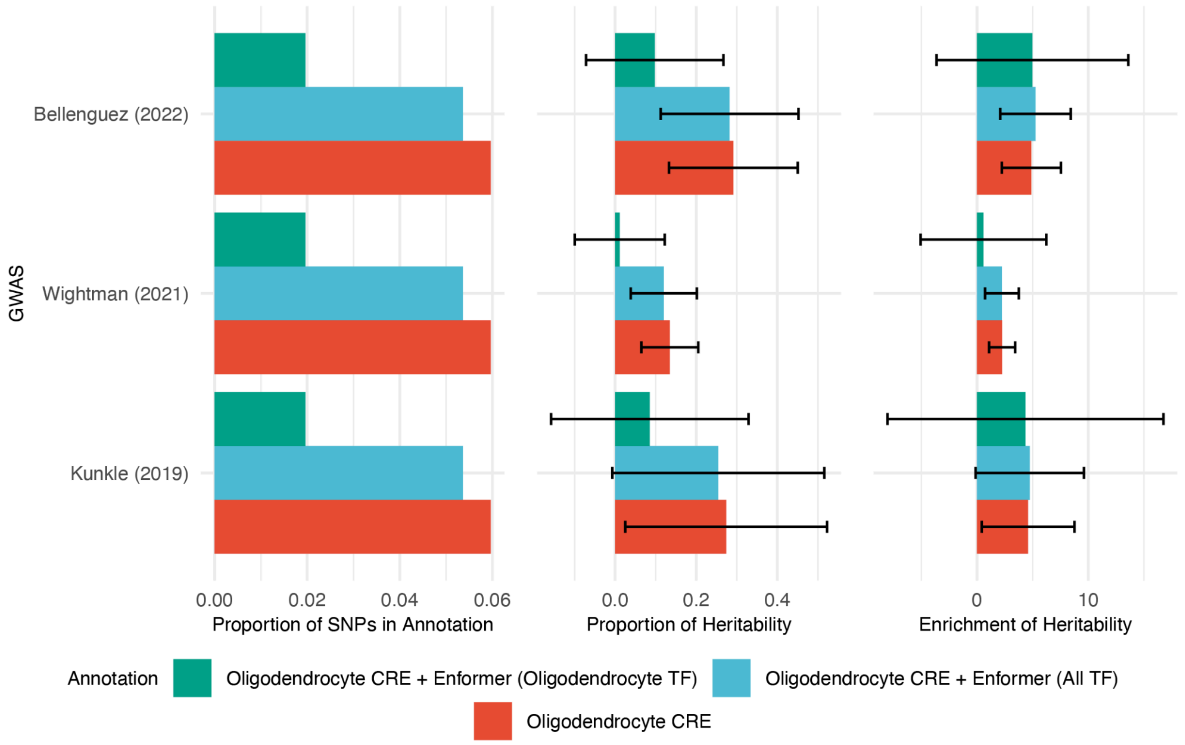


Supplementary Figure 3: Comparison of enrichment and proportion of heritability for cell-type specific TF (CTF) versus all TF (ATF) annotations at different z-score thresholds for oligodendrocytes for AD GWAS.

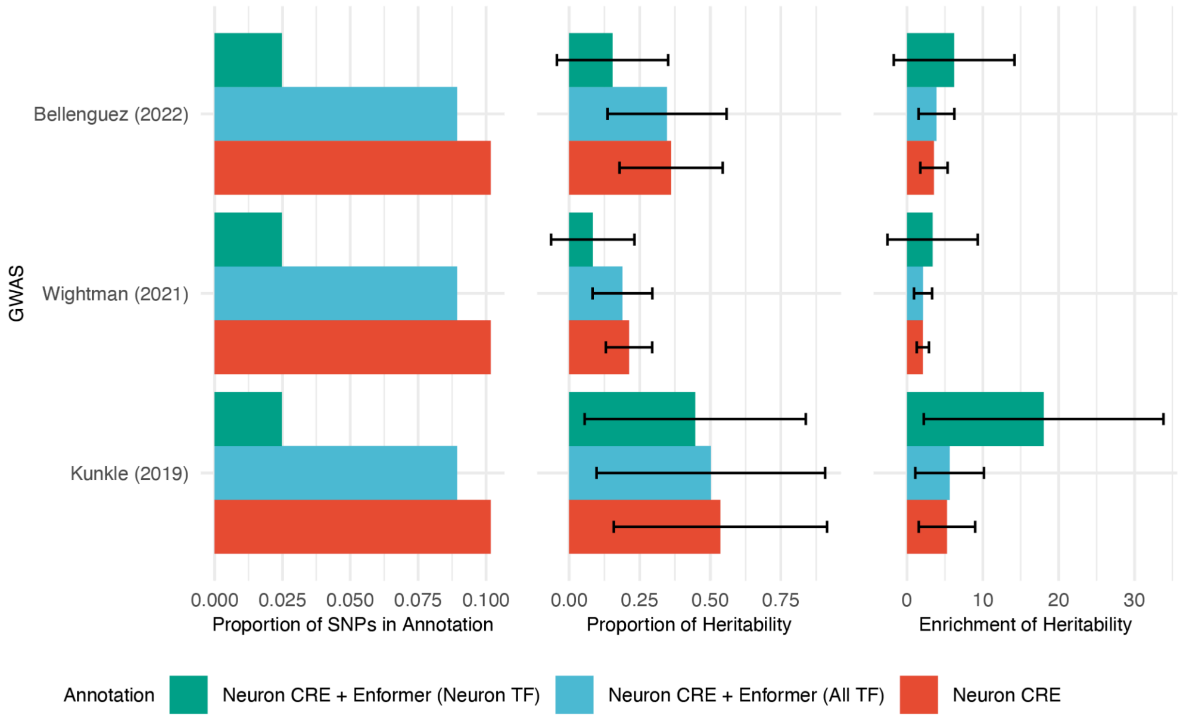


Supplementary Figure 4: Comparison of enrichment and proportion of heritability for cell-type specific TF (CTF) versus all TF (ATF) annotations at different z-score thresholds for neurons for AD GWAS.

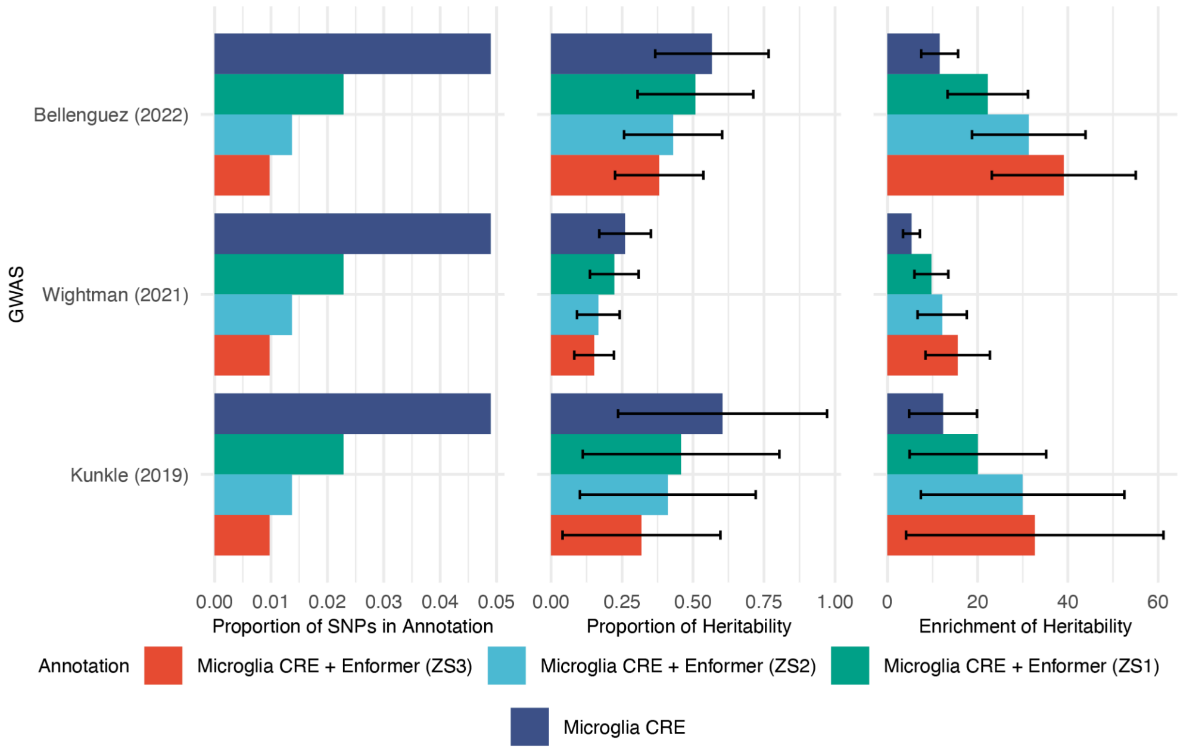


Supplementary Figure 5: Relationship between z-score threshold stringency for microglia TF disruption and both enrichment of heritability (e) and proportion of heritability (p) for AD GWAS.

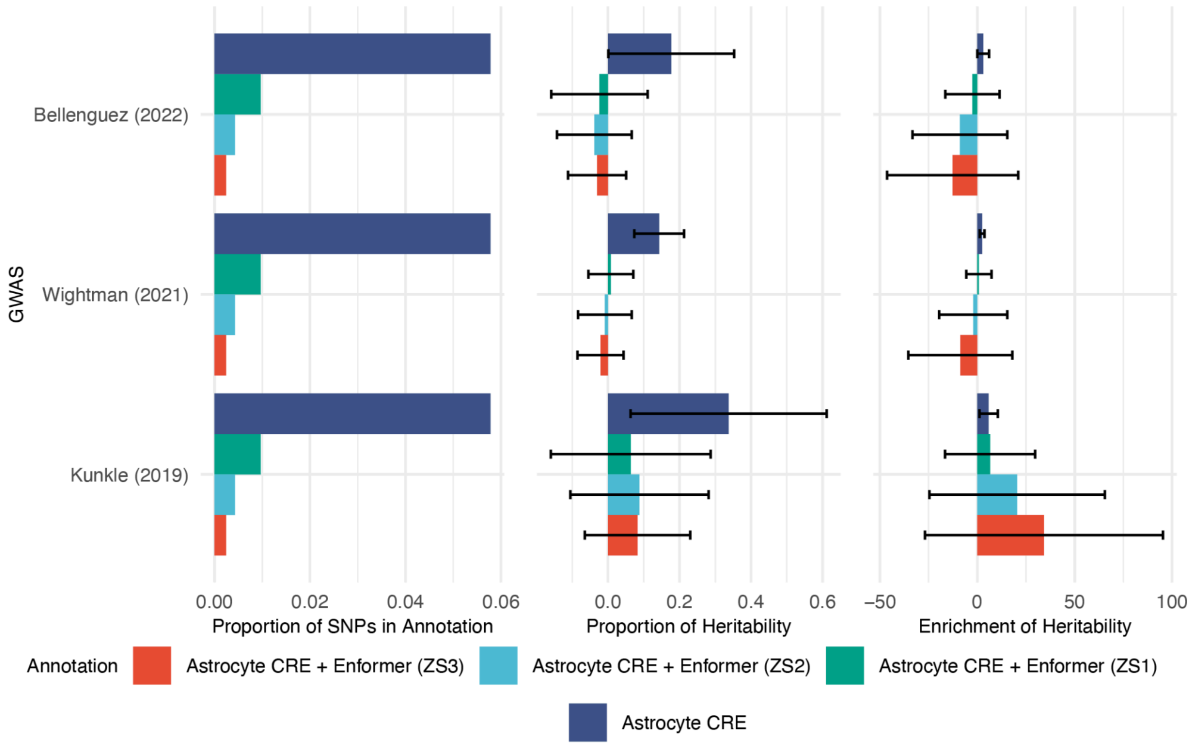


Supplementary Figure 6: Relationship between z-score threshold stringency for astrocyte TF disruption and both enrichment of heritability (e) and proportion of heritability (p) for AD GWAS.

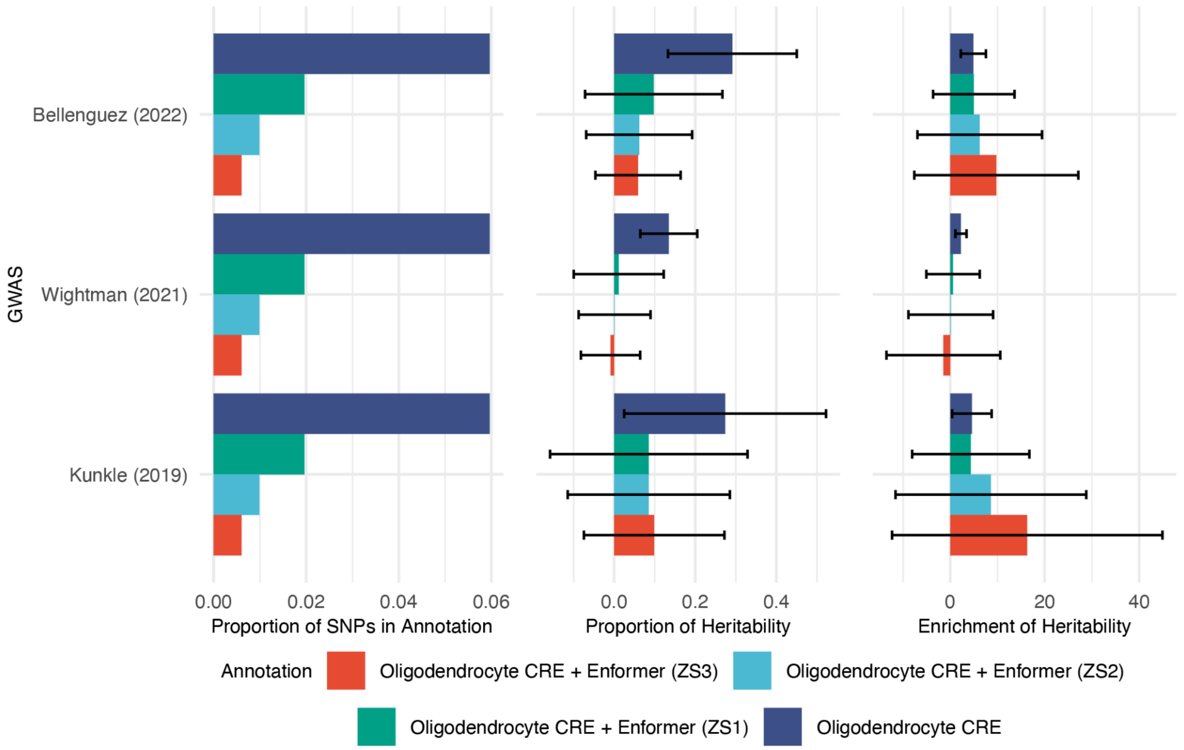


Supplementary Figure 7: Relationship between z-score threshold stringency for oligodendrocyte TF disruption and both enrichment of heritability (e) and proportion of heritability (p) for AD GWAS.

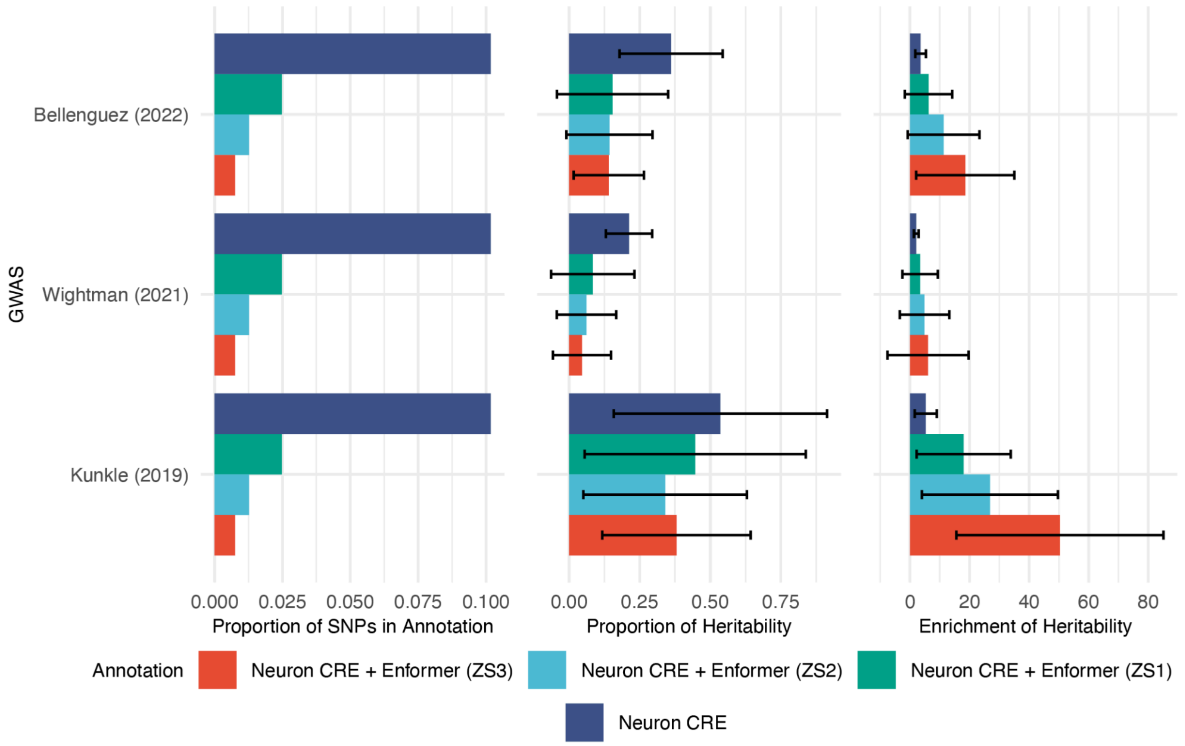


Supplementary Figure 8: Relationship between z-score threshold stringency for neuronal TF disruption and both enrichment of heritability (e) and proportion of heritability (p) for AD GWAS.

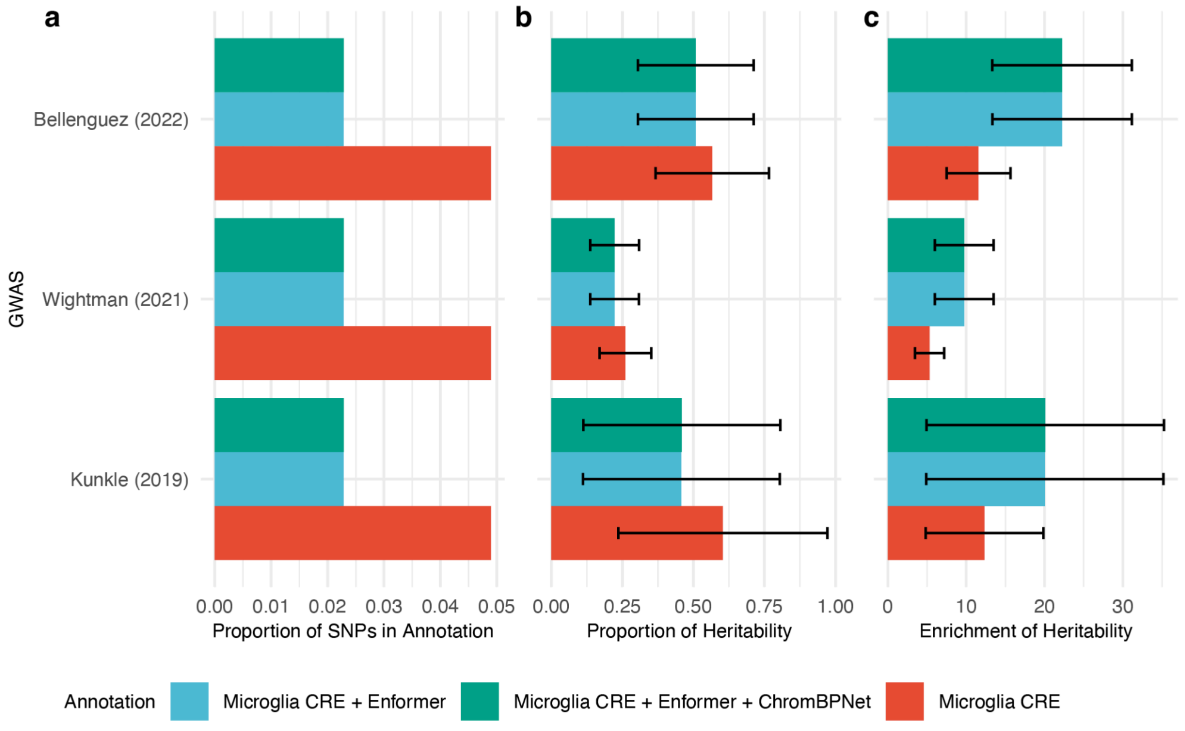


Supplementary Figure 9: Comparison of enrichment and proportion of heritability within microglia CRE, microglia CRE + Enformer, and microglia CRE + Enformer + ChromBPNet annotations for AD GWAS.

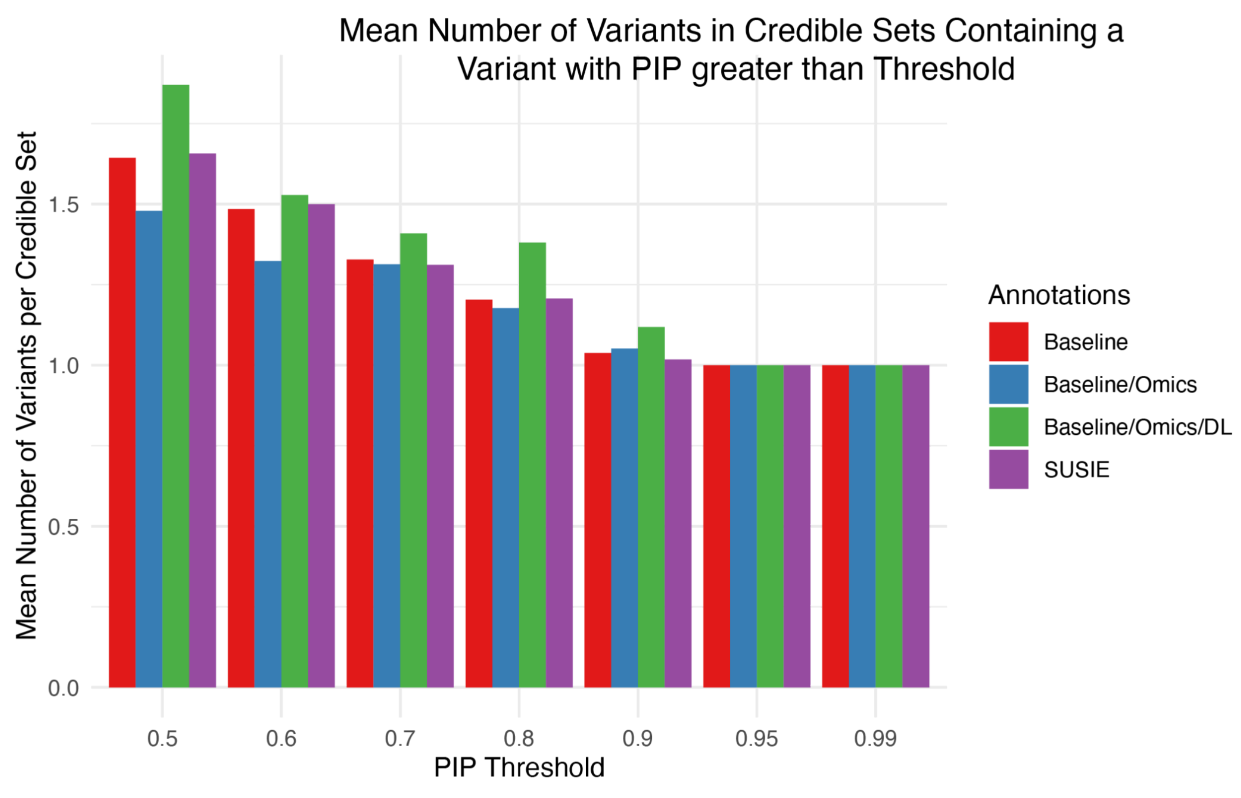


Supplementary Figure 10: Distribution of credible set sizes from different fine-mapping approaches, showing larger mean set size (n=1.77) for Baseline/Omics/DL versus SuSiE (n=1.51).

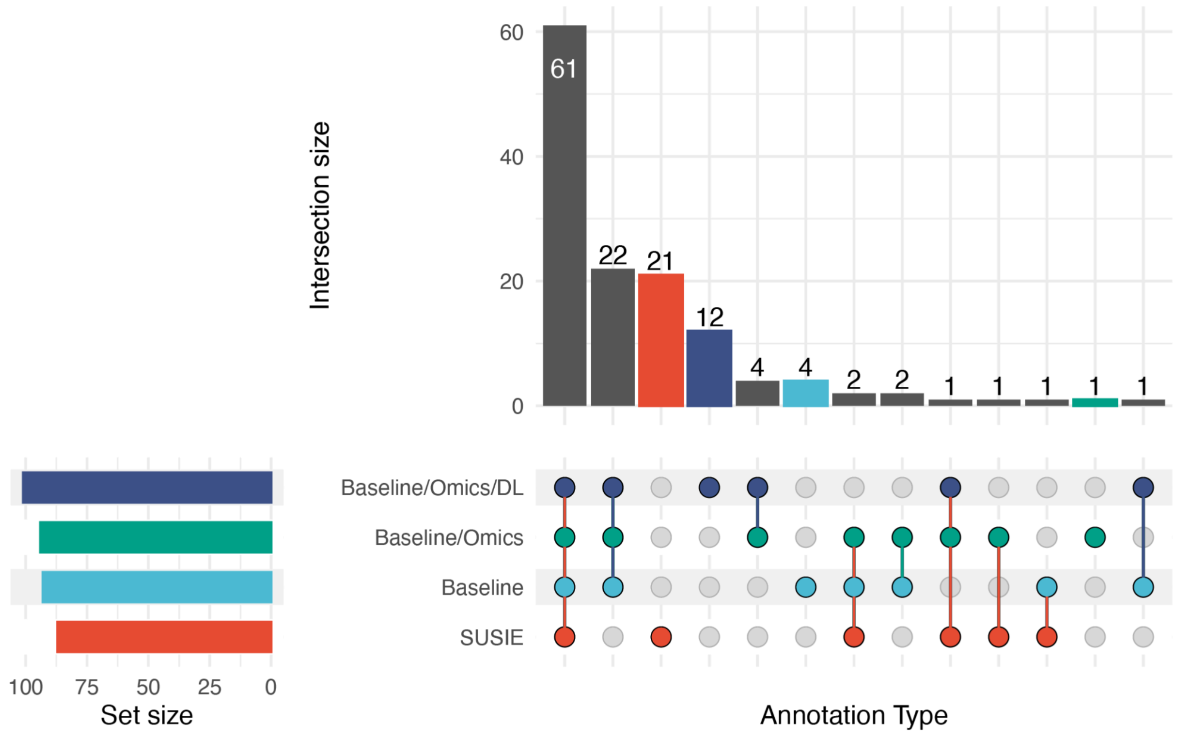


Supplementary Figure 11: UpsetR plot showing the distribution of 138 nominally significant SNPs (PIP>0.5) across four fine-mapping approaches.

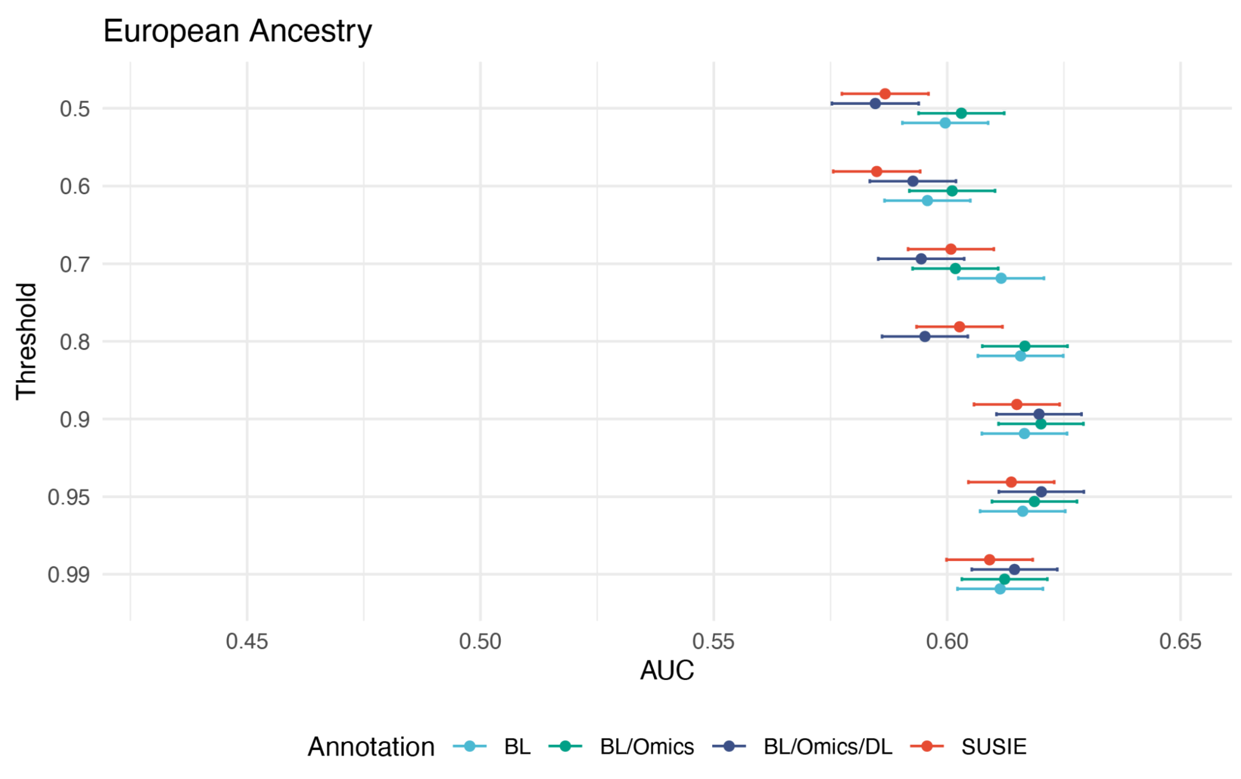


Supplementary Figure 12: Comparison of AUC values for PRS based on different fine-mapping approaches for the ADSP individuals of European ancestry at varying PIP thresholds.

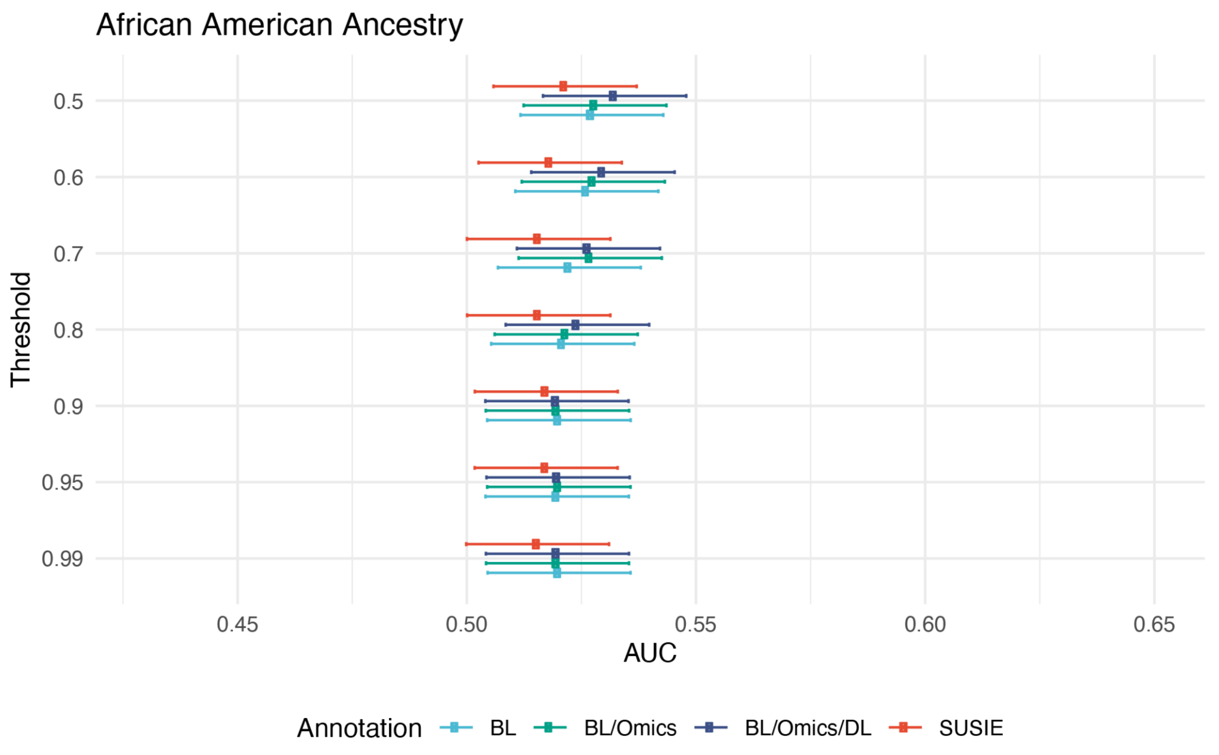


Supplementary Figure 13: Comparison of AUC values for PRS based on different fine-mapping approaches for the ADSP individuals of African American ancestry at varying PIP thresholds.

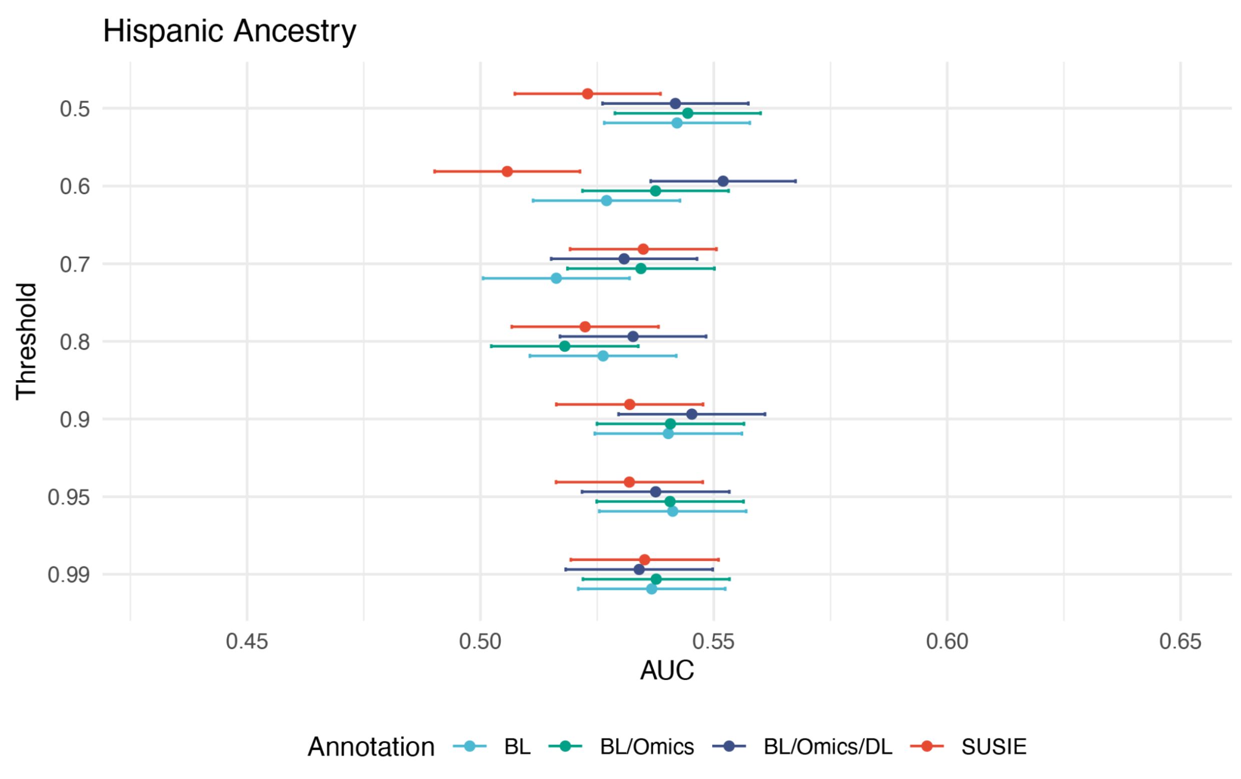


Supplementary Figure 14: Comparison of AUC values for PRS based on different fine-mapping approaches for the ADSP individuals of Hispanic ancestry at varying PIP thresholds.

### LocusZoom Plots

#### Locus 1

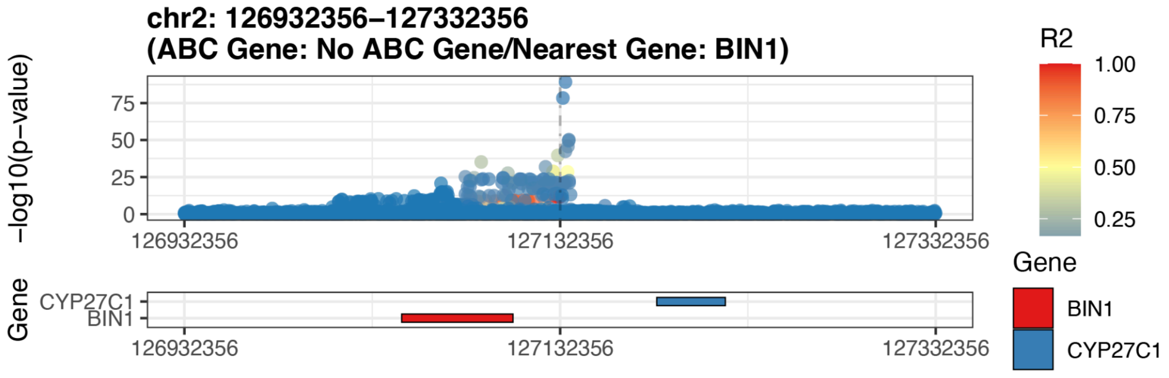


Supplementary Figure 15: Gene locus plot for locus 1

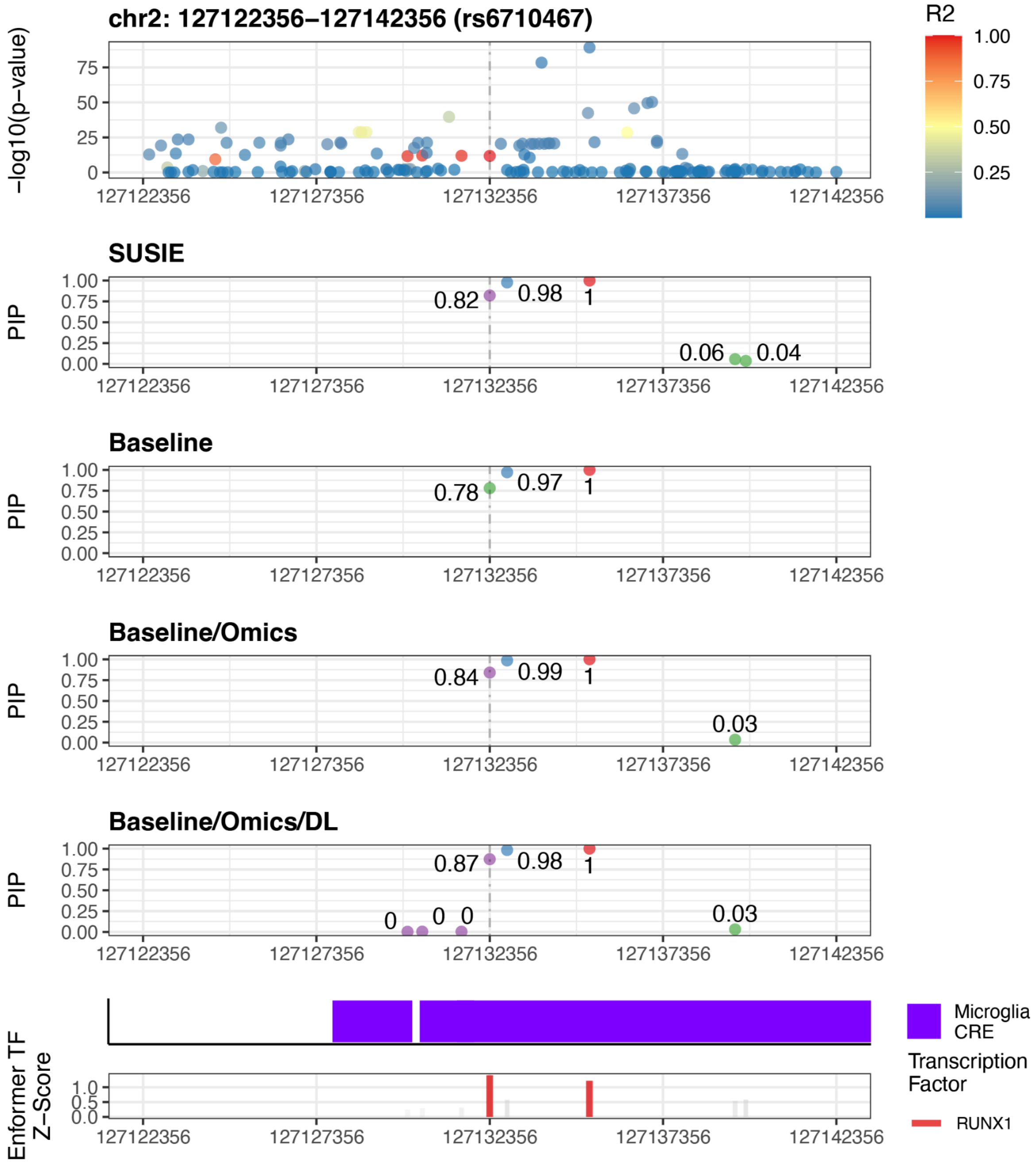


Supplementary Figure 16: Zoomed-in locus plot for locus 1

Locus 2

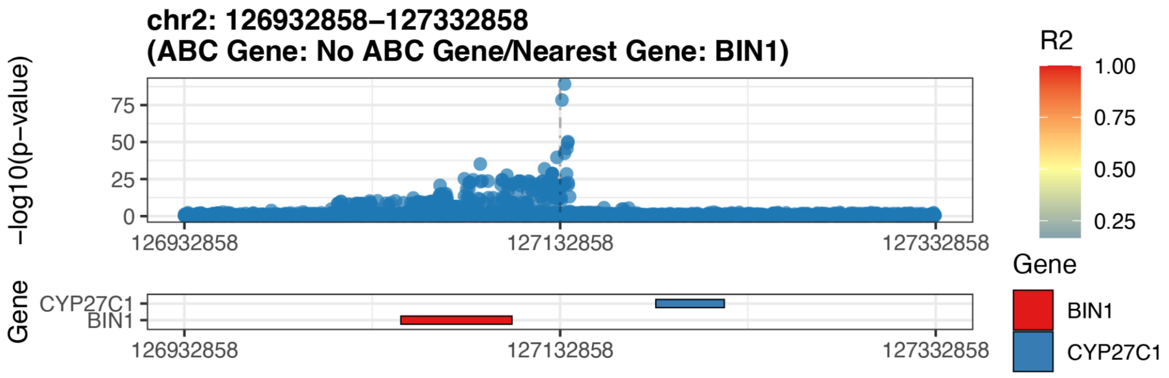


Supplementary Figure 17: Gene locus plot for locus 2

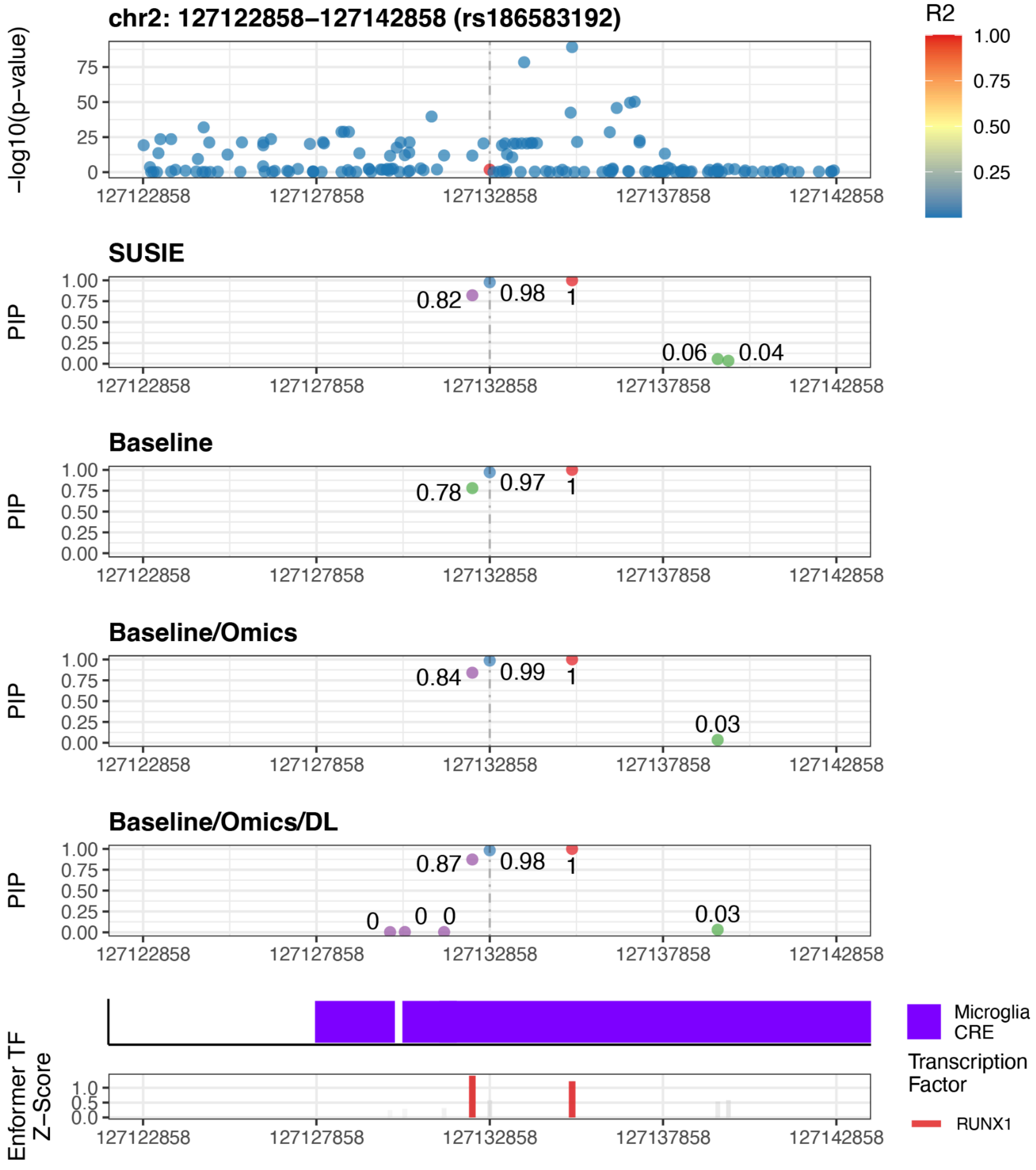


Supplementary Figure 18: Zoomed-in locus plot for locus 2

Locus 3

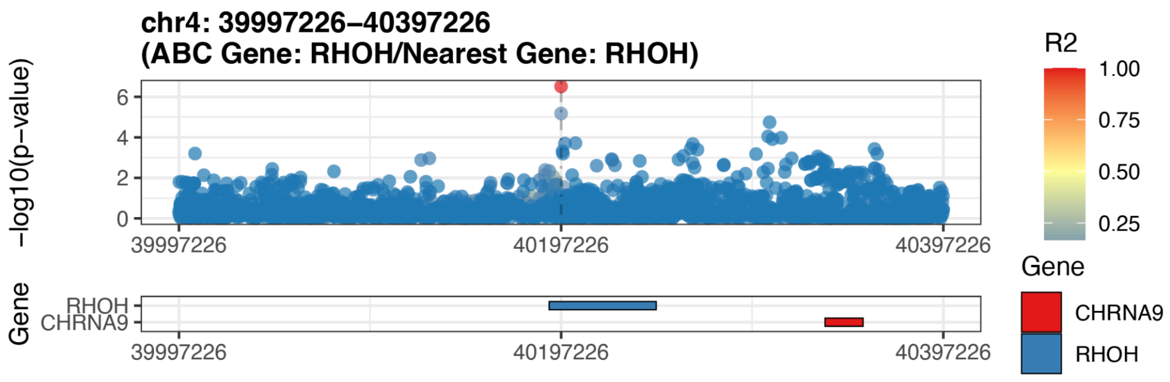


Supplementary Figure 19: Gene locus plot for locus 3

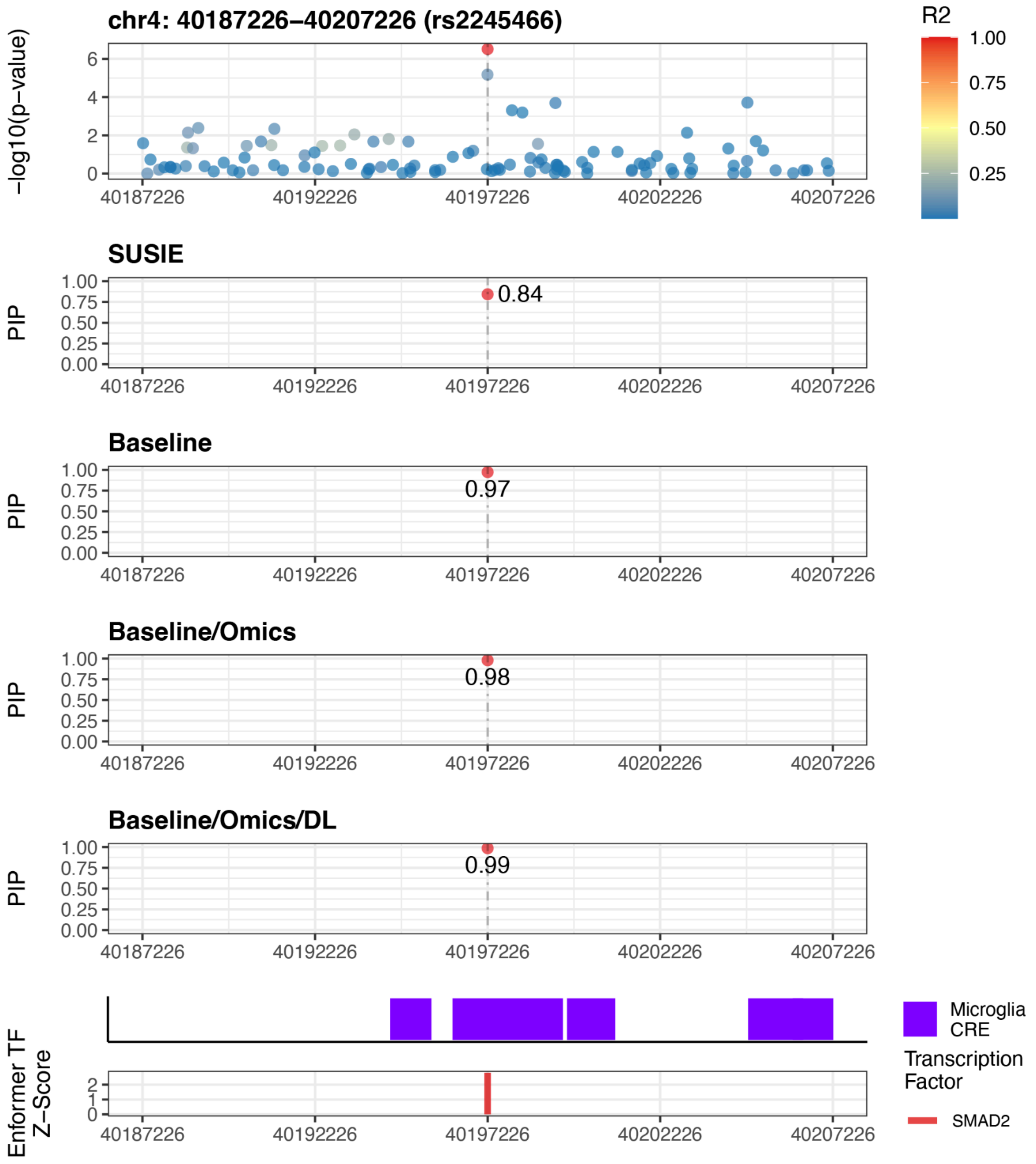


Supplementary Figure 20: Zoomed-in locus plot for locus 3

#### Locus 4

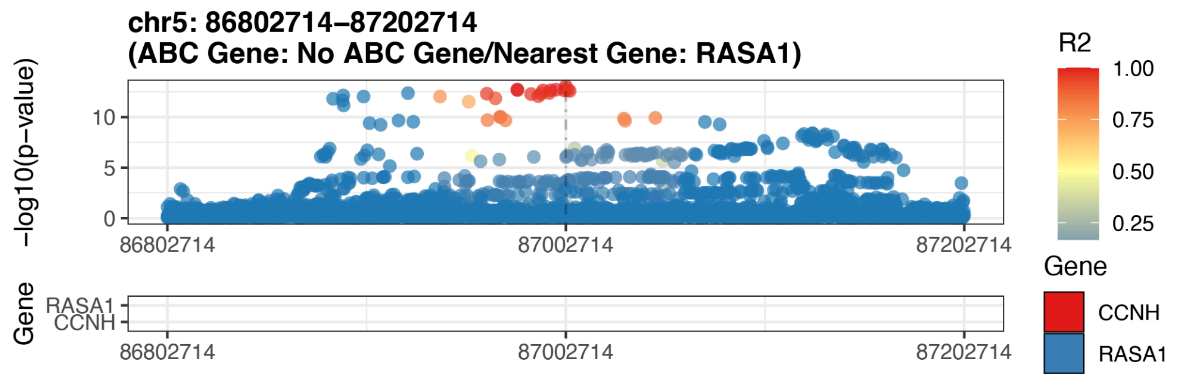


Supplementary Figure 21: Gene locus plot for locus 4

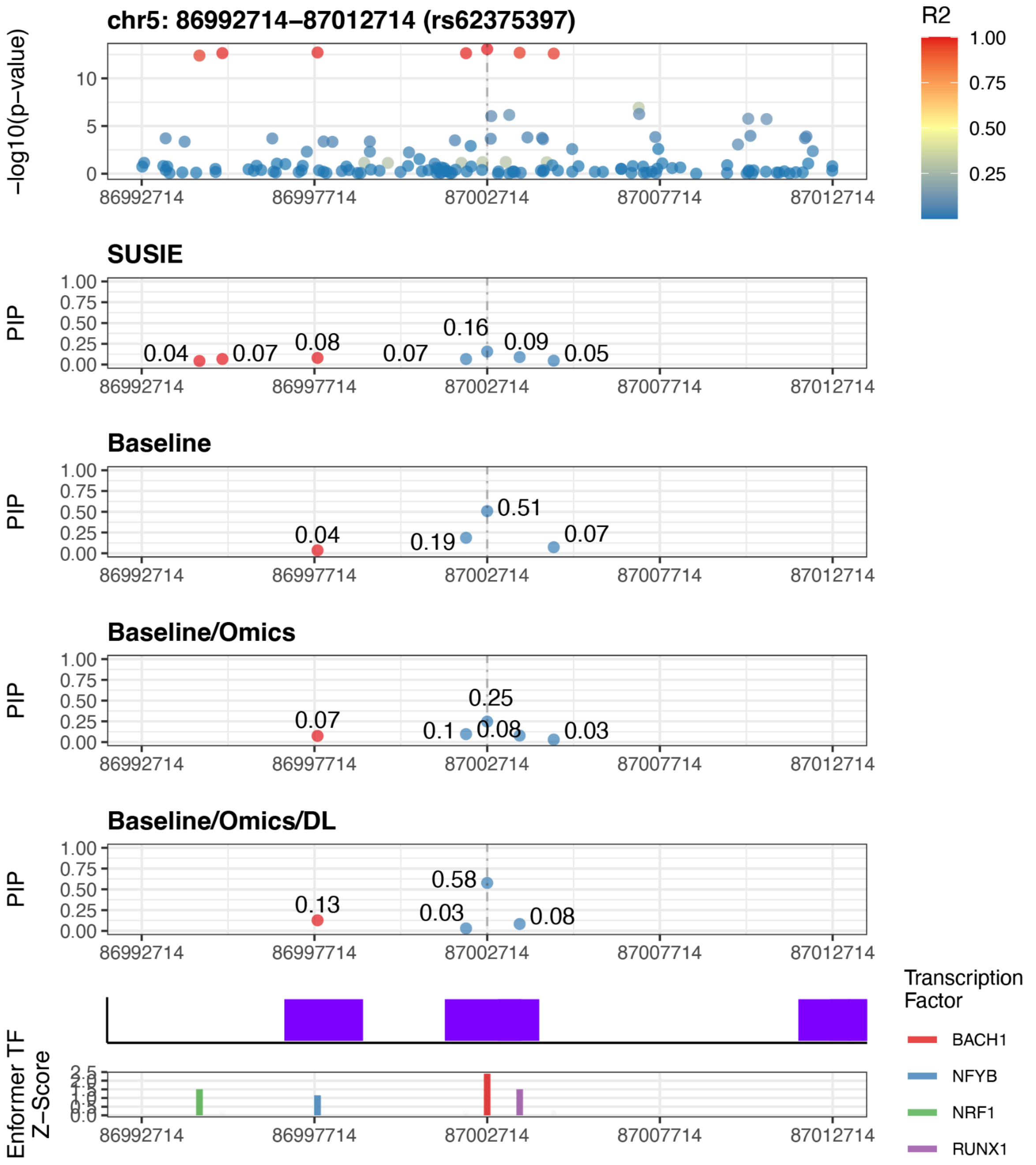


Supplementary Figure 22: Zoomed-in locus plot for locus 4

#### Locus 5

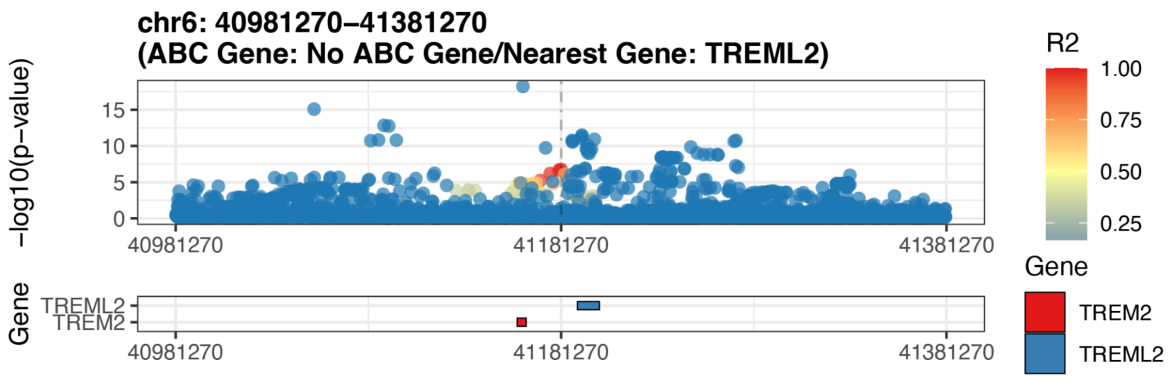


Supplementary Figure 23: Gene locus plot for locus 5

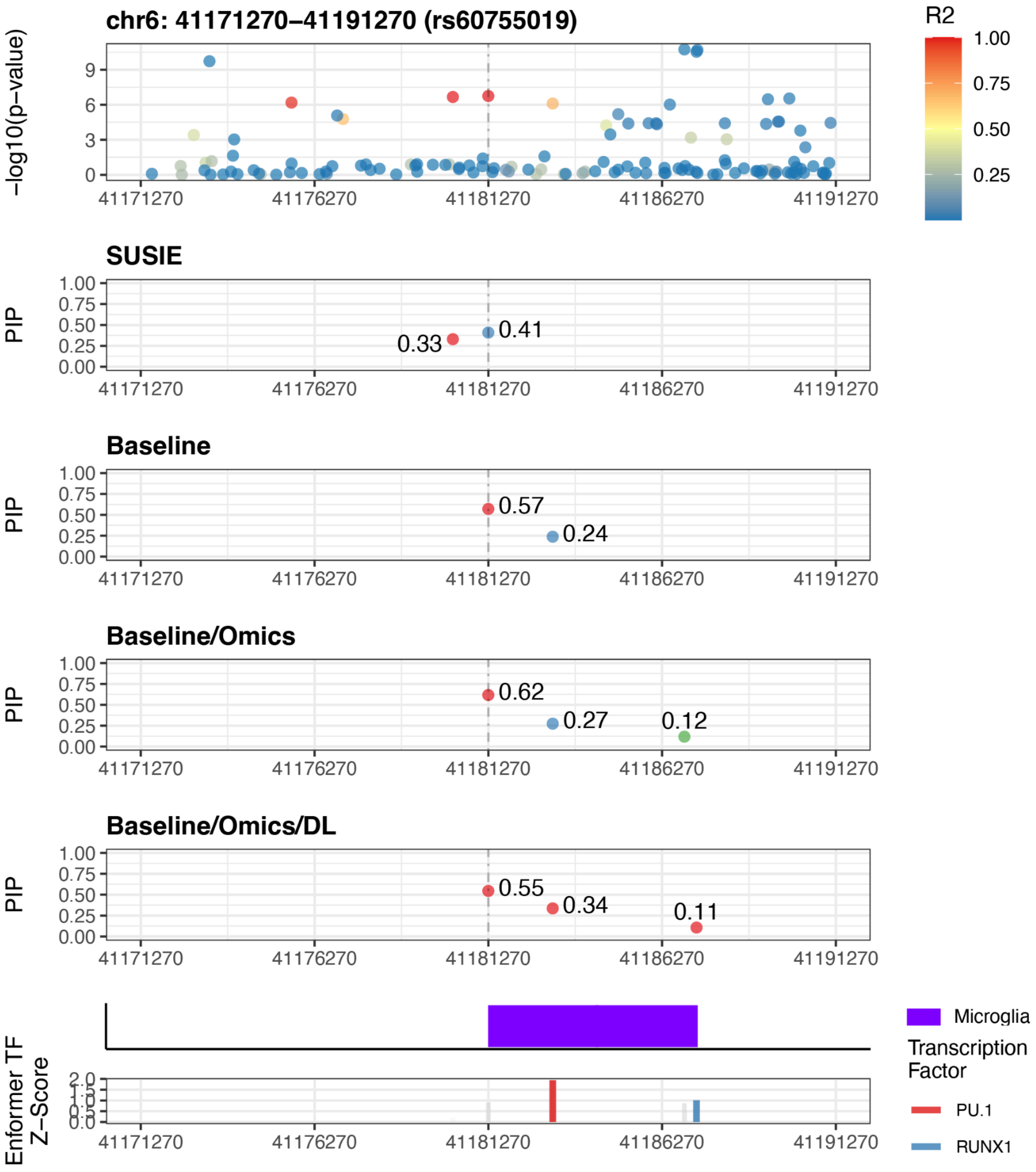


Supplementary Figure 24: Zoomed-in locus plot for locus 5

#### Locus 6

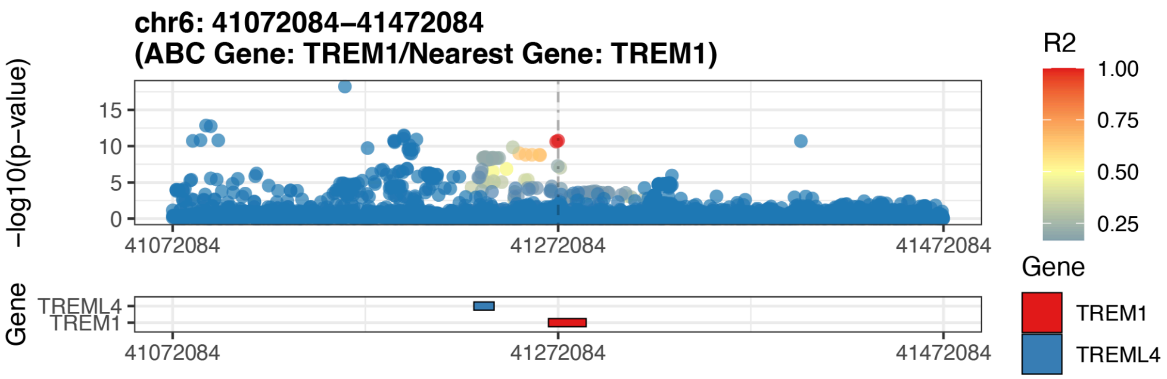


Supplementary Figure 25: Gene locus plot for locus 6

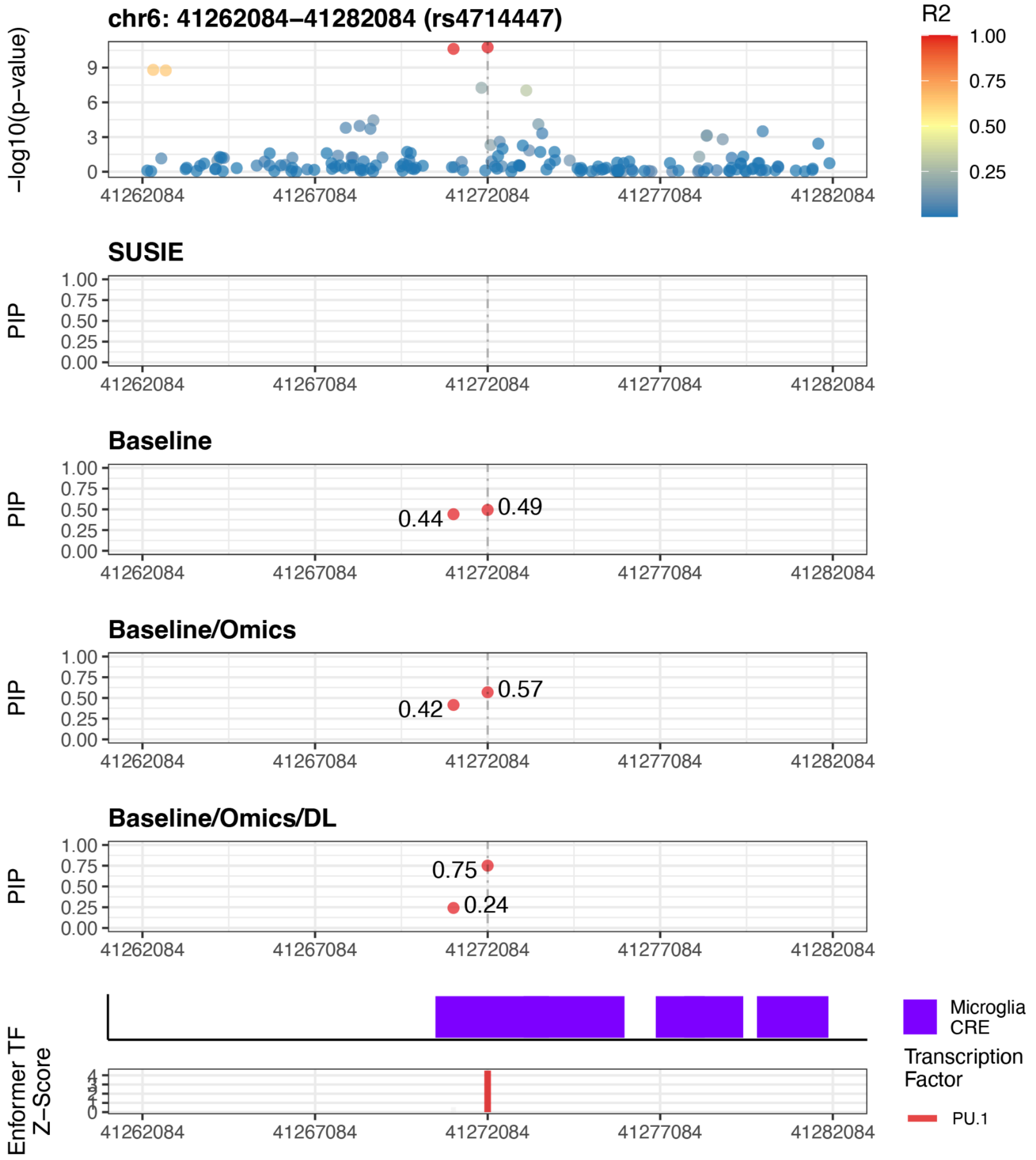


Supplementary Figure 26: Zoomed-in locus plot for locus 6

#### Locus 7

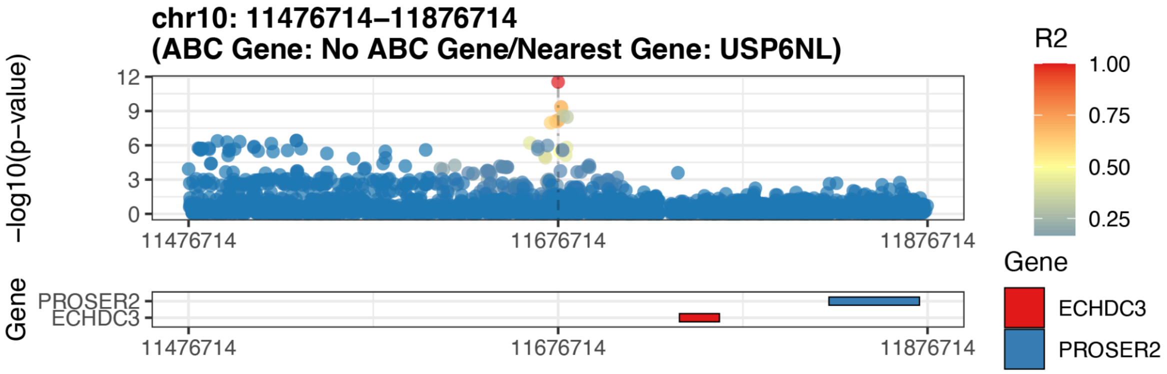


Supplementary Figure 27: Gene locus plot for locus 7

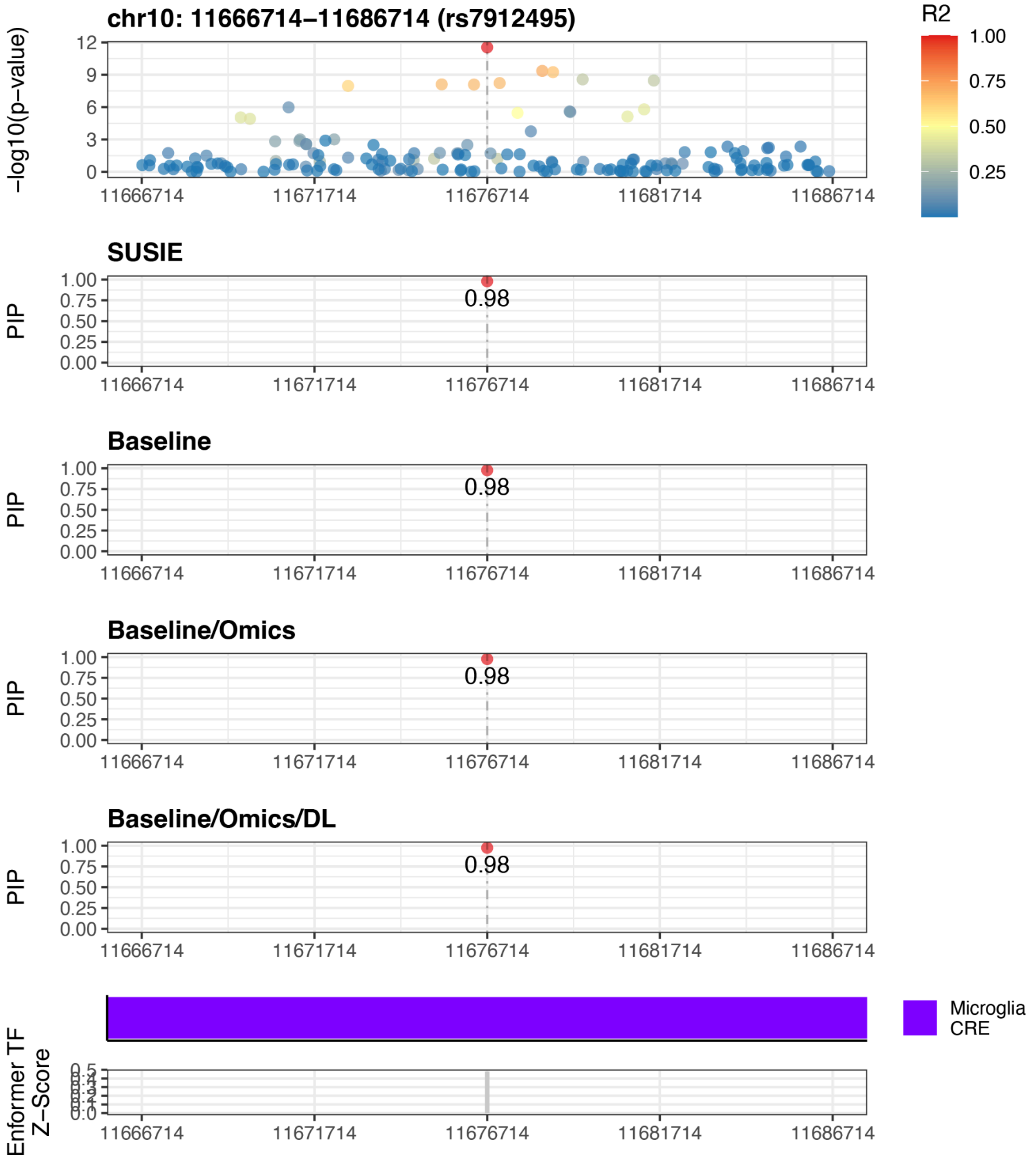


Supplementary Figure 28: Zoomed-in locus plot for locus 7

#### Locus 8

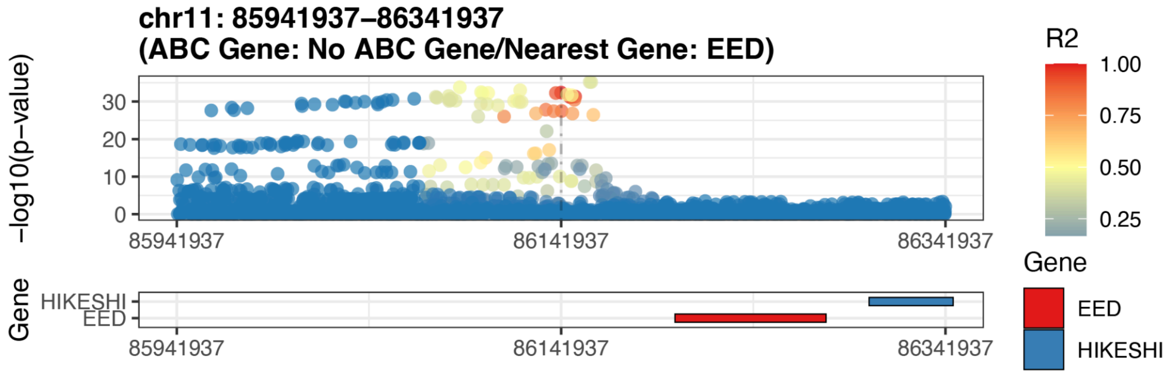


Supplementary Figure 29: Gene locus plot for locus 8

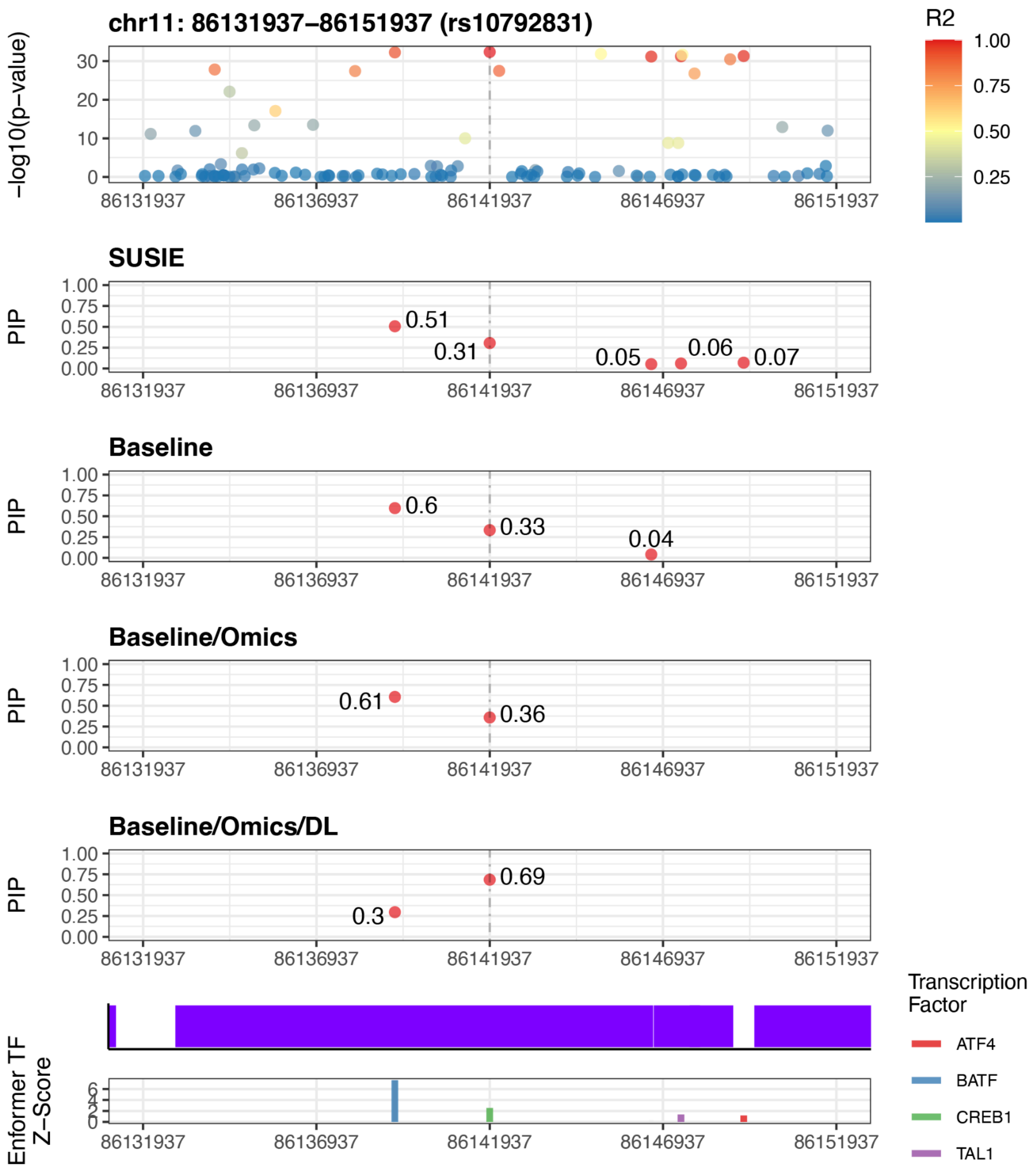


Supplementary Figure 30: Zoomed-in locus plot for locus 8

#### Locus 9

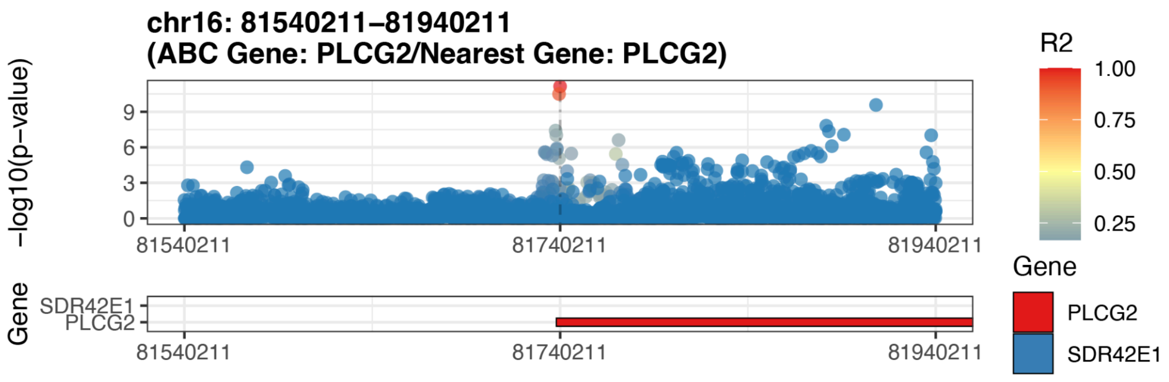


Supplementary Figure 31: Gene locus plot for locus 9

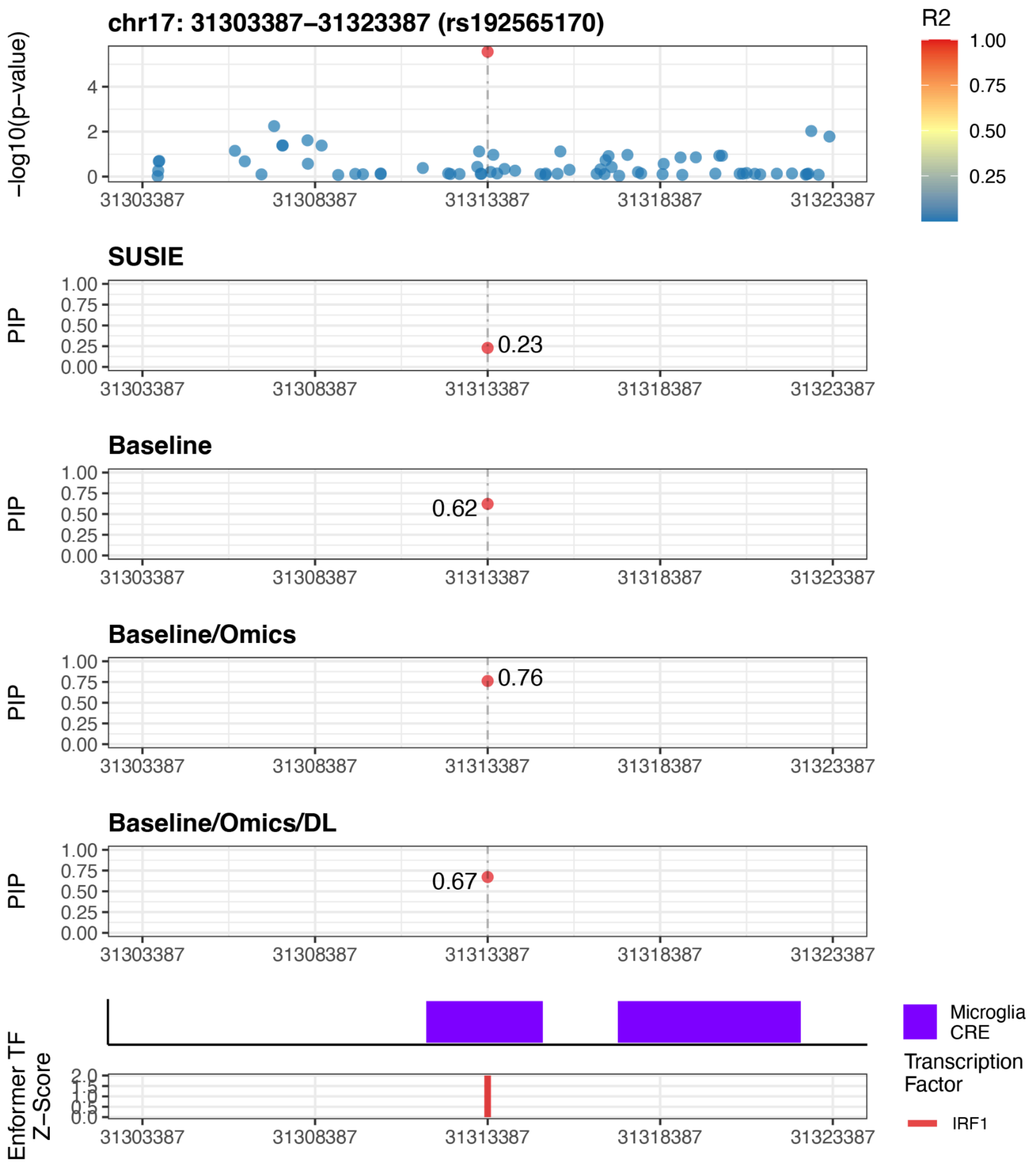


Supplementary Figure 32: Zoomed-in locus plot for locus 9

Locus 10

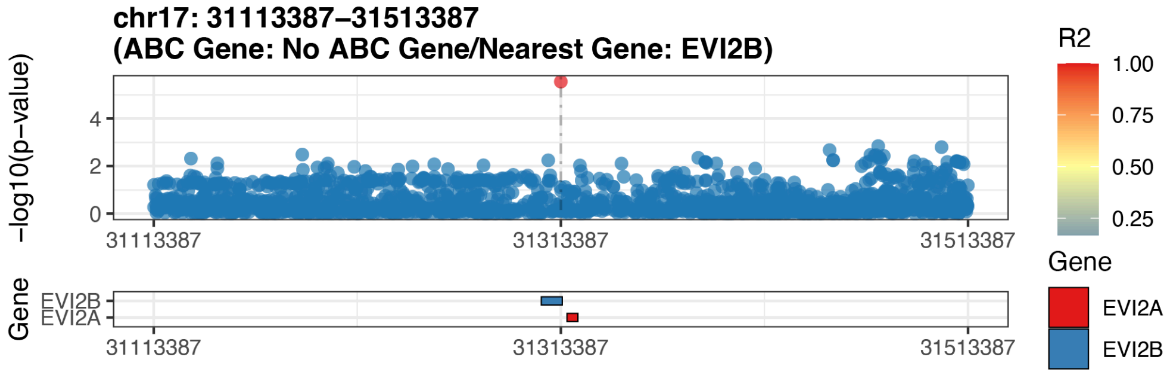


Supplementary Figure 33: Gene locus plot for locus 10

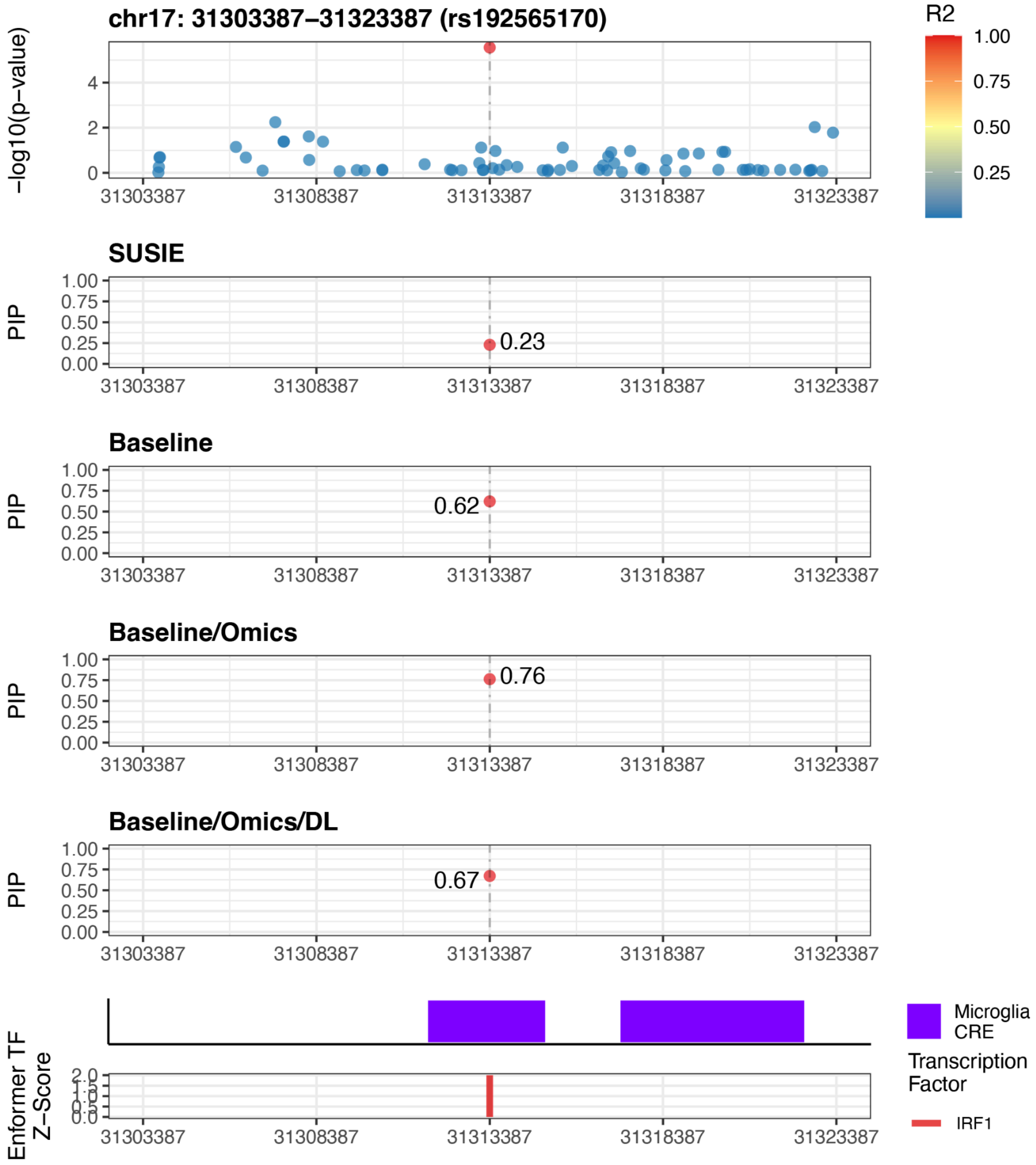


Supplementary Figure 34: Zoomed-in locus plot for locus 10

Locus 11

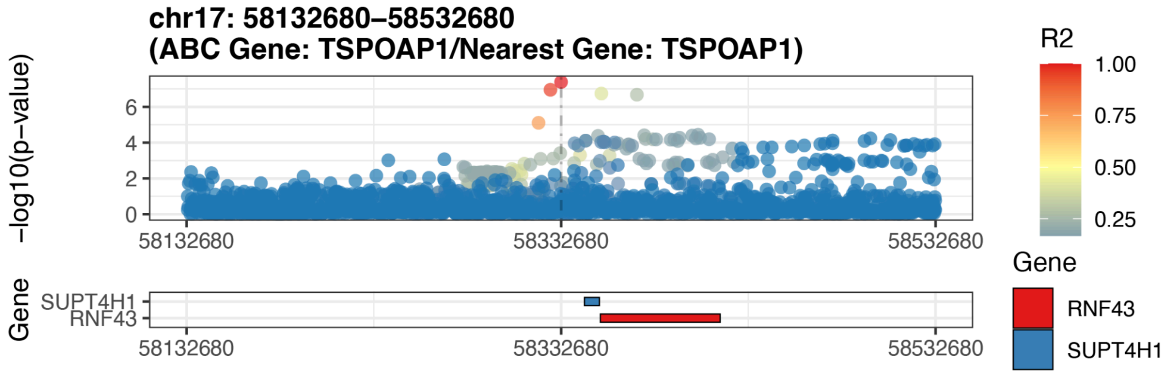


Supplementary Figure 35: Gene locus plot for locus 11

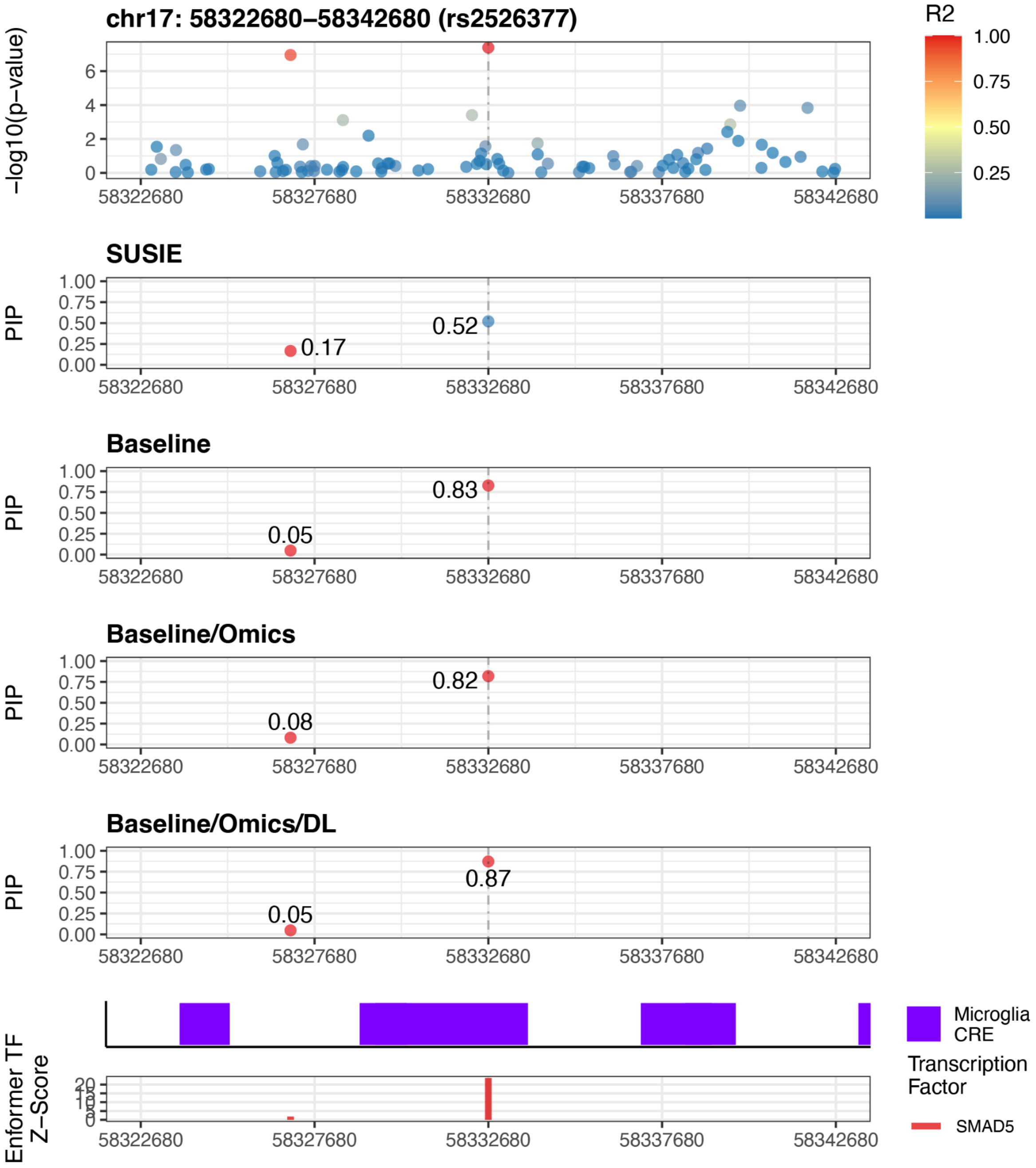


Supplementary Figure 36: Zoomed-in locus plot for locus 11

#### Locus 12

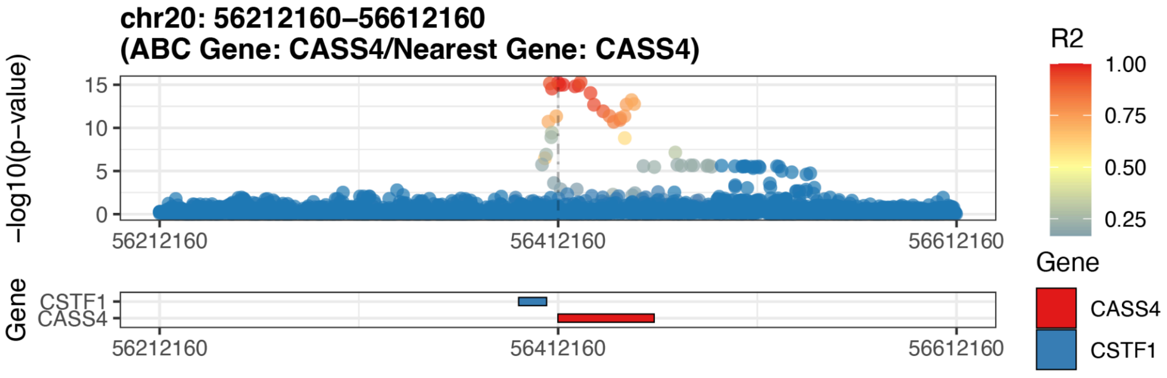


Supplementary Figure 37: Gene locus plot for locus 12

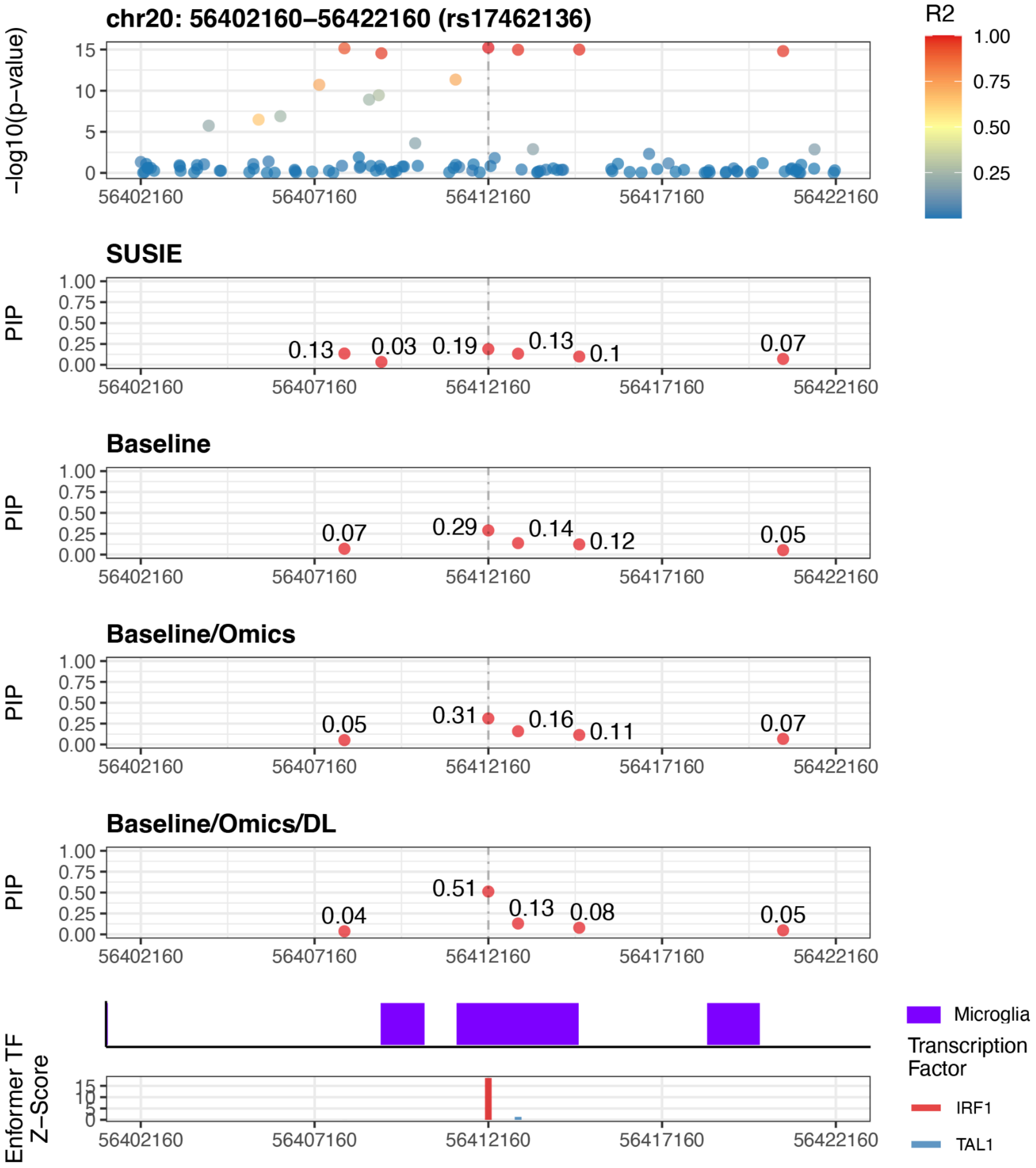


Supplementary Figure 38: Zoomed-in locus plot for locus 12

#### Supplementary Tables

| Annotation Category | Number of Annotations | Number of Cell Types | Number of Assays/CRE Regions | Thresholds |
|---|---|---|---|---|
| Baseline | 187 | N/A | N/A | N/A |
| Roadmap | 635 | 127 | 5 | N/A |
| Brain Omics | 40 | 4 | 10 | N/A |
| DeepSea (Beluga) | 5896 | 233 | 37 | Z score > 1<br>Z score > 2<br>Z score > 3 |
| Enformer | 13518 | 505 | 35 | Z score > 1<br>Z score > 2<br>Z score > 3 |
| Enformer + Brain Omics | 240 | 4 | 1 | Z score > 1<br>Z score > 2<br>Z score > 3 |

Supplementary Table 1: Counts of Annotations by Category

| Cell Type | Transcription Factors |
|---|---|
| Astrocytes | MEIS2, NFIB, TGIF1, EMX2, LHX2, RFX2, RFX4, RORA, RORB, SOX1, SOX2, SOX21, SOX9, NFATC4, SOX5, POU3F2, POU3F3, POU3F4, FOXG1, FOXO1, SP5 |
| Microglia | ETS2, FLI1, SPI1, IRF8, PRDM1, CEBPA, CEBPB, CEBPD, CEBPE, KLF11, KLF2, TFEC, BACH1, BATF, BATF3, RUNX1, RUNX2, RUNX3, LYL1, TAL1, ELK3, MEF2C, CREB3L2, ATF4, MAFB, SALL1, eGFP-SALL1, SMAD5, MEF2A, MEF2B, SMAD2, USF1, STAT3, eGFP-MAFG, NFYB, NRF1, CREB1, IRF1, SOX9 |
| Oligodendrocytes | SOX10, SOX13, SOX21, SOX3, SOX6, SOX8, NHLH2, NFIX, SP7, NFE2, E2F1, CREB5, POU3F3, MYCN |
| Neuron | ASCL1, ASCL5, BHLHA15, BHLHE22, NEUROD1, NEUROD2, NEUROD6, TWIST2, EGR4, SP8, SP9, MEIS3, PKNOX2, MSC, KLF5, KLF8, TBR1, HLF |

Supplementary Table 2: Transcription factors for each cell type

Supplementary Table 3

supplementary_table_3.tsv

Supplementary Table 4

supplementary_table_4.tsv

| Annotation | Ancestry | AUC (SE) | SE | PIP Threshold |
|---|---|---|---|---|
| SUSIE | EUR | 0.615 | 0.00468 | 0.9 |
| Baseline | EUR | 0.617 | 0.00465 | 0.9 |
| Baseline/Omics | EUR | 0.620 | 0.00465 | 0.9 |
| Baseline/Omics/DL | EUR | 0.620 | 0.00465 | 0.95 |
| SUSIE | AFR | 0.521 | 0.00797 | 0.5 |
| Baseline | AFR | 0.527 | 0.00795 | 0.5 |
| Baseline/Omics | AFR | 0.528 | 0.00795 | 0.5 |
| Baseline/Omics/DL | AFR | 0.532 | 0.00797 | 0.5 |
| SUSIE | AMR | 0.535 | 0.00807 | 0.99 |
| Baseline | AMR | 0.542 | 0.00795 | 0.5 |
| Baseline/Omics | AMR | 0.544 | 0.00796 | 0.5 |
| Baseline/Omics/DL | AMR | 0.552 | 0.00792 | 0.6 |

Supplementary Table 5: Top PRS AUC for each ancestry across the 4 fine-mapping strategies
